# The Burden of Malignant Skin Melanoma in China: From 1990 to 2021 with Estimation to 2036

**DOI:** 10.1101/2025.08.11.25333476

**Authors:** Chenyu Zhao, Keqiang Ma, Zhaoqi Wu, Ruihan Zhang, Yiting He, Martin Gluchman, Yiting Wang, Hui Wang

## Abstract

**Background:** Malignant skin melanoma (MSM), a highly aggressive skin cancer from melanocytes, has a higher mortality rate than other skin cancers due to its metastatic nature. Despite advancements in diagnostics and treatment, melanoma remains a significant health concern in China.

**Methods:** Data from the Global Burden of Disease (GBD) 2021 database was utilized to analyze MSM trends in China from 1990 to 2021. Statistical analyses, including Joinpoint regression and decomposition analysis, were employed to examine changes in incidence and mortality. The ARIMA model was used to project age-standardized incidence and mortality rates through 2036.

**Results:** In 2021, China reported approximately 13,437 new cases of MSM, with a total patient population of 81,219 and 5,373 MSM-related deaths. Between 1990 and 2021, the incidence and prevalence rates of MSM significantly increased, while mortality rates showed a slight decline. The highest incidence rate was observed in the 55-59 age group, with males facing notably higher risks than females. Joinpoint regression analysis indicated rapid Age-Standardized Incidence Rate (ASIR) growth from 2004 to 2012, and decomposition analysis identified population aging as the primary driving factor. ARIMA forecasts suggest that by 2036, ASIR will reach 0.88 for males and 0.71 for females, while Age-Standardized Death Rate (ASDR) will stabilize or decline.

**Conclusion:** The MSM burden in China has intensified, with increasing incidence and prevalence rates over time. Although mortality rates have declined slightly, the impact of melanoma on public health remains significant.

## 1. Introduction

Malignant melanoma is a highly aggressive type of skin cancer, originating from melanocytes, the pigment-producing cells in the skin [1]. It is considered the most severe form of skin cancer. Prolonged exposure to ultraviolet radiation is the primary etiological factor for this disease, as it leads to DNA mutations in melanocytes [2]. Genetic factors, such as BRAF and NRAS mutations, are directly associated with a significant number of melanoma cases [3]. Furthermore, immunocompromised individuals, including those with HIV or organ transplant recipients on long-term immunosuppressive therapy, are at increased risk due to their reduced ability to identify and eliminate mutated melanocytes [4]. Although melanoma accounts for a small proportion of skin cancer cases, its high invasiveness and propensity for distant metastasis result in a much higher mortality rate compared to other skin cancers [5].

Over the past two decades, diagnostic and therapeutic approaches for malignant melanoma have progressively improved. A combination of dermoscopy, imaging, and genetic testing has systematized the recognition of melanoma’s warning signs, utilizing the ABCDE rule [6]. In early-stage melanoma, surgical excision of the primary lesion and a margin of healthy surrounding tissue remains the mainstay of treatment [7][8]. For advanced melanoma, immunotherapies such as PD-1 and CTLA-4 inhibitors have significantly improved survival rates [9]. Radiation therapy is frequently applied in advanced or metastatic cases, particularly those with metastases to the brain, bones, or other inoperable sites [10]. Chemotherapy, although less commenly used, is an option when other treatments have failed, serving as an adjunctive therapeutic strategy [11].

However, outcomes for melanoma patients in China are less favorable compared to those in Western countries, likely due to the predominance of acral and mucosal melanoma subtypes in the Chinese population [12]. These subtypes are often more challenging to treat [13]. Although many immunotherapies present promising expectations the Chinese Society of Clinical Oncology (CSCO) has only recently incorporated these therapies into its guidelines for adjunctive treatment [14]. The vast amount of data supporting the use of immunotherapy is derived from studies on Caucasian populations, potentially limiting the treatment benefits for Chinese patients. Moreover, Chinese melanoma patients often face barriers to obtain comprehensive and accurate information regarding their disease.

A systematic analysis of major databases reveals that epidemiological research on melanoma in China remains insufficient. This highlights the need for increased attention and support to better address the burden of melanoma in the region. In this study, data from the GBD 2021 database were used to describe the trends of malignant skin melanoma burden in China from 1990 to 2021. In addition, we have projected the MSM incidence and mortality rates in in China by 2036, providing a more comprehensive reference value.

## 2. Methods

GBD 2021 provides a comprehensive analysis of global health trends, encompassing 371 diseases and injuries across 204 countries and territories [15]. We used keywords such as “skin cancer”, “melanoma”, “incidence”, “prevalence”, “mortality”, “DALY”, and “epidemiology” to search PubMed, ScienceDirect, Web of Science, and official websites of the Chinese Government and other institutions for studies and reports on MSM in China published in English or Chinese until October 1, 2024. While previous studies have provided comprehensive national-level analyses of the skin cancer burden in China, consistent and focused trend analysis of MSM have been lacking [16]. In this study, we extracted the latest data from GBD 2021 to analyze the disease burden of MSM in China. The analysis includes all gender and age groups provided by the GBD database. The data acquisition process is detailed in previous studies, and all acquired data underwent rigorous manual quality control (by C.Z).

### 2.1 Data sources and measures

The International Classification of Diseases 10th revision (ICD-10) codes for MSM are C43.0 - C43.9 [15]. Mortality data for MSM in the GBD 2021 China study were mainly sourced from national disease surveillance point systems, death cause registration reporting systems, and maternal and child health surveillance websites. Data on non-fatal outcomes of MSM were collected from population-based surveys and census data, as well as other published and unpublished studies and reports. Only data that met quality control criteria were included in the modelling estimation [17]. The Cause of Death Ensemble Model (CODEm) was employed to estimate cause-specific mortality and years of life lost (YLLs), while DisMod-MR 2.1 was utilized to assess disease burden, including incidence and years lived with disability (YLDs) [18]. Disability-Adjusted Life Years (DALYs) represents the total health loss due to diseases and injuries. It is the sum of YLDs and YLLs. All data, along with their corresponding 95% uncertainty intervals (UIs), were derived from cancer registries, published literature, surveillance data, census data, and other data sources, stratified by location, sex, and age group [15].

### 2.2 Joinpoint regression model

The Joinpoint regression model is a statistical method used primarily to analyze trends over time. It allows for the calculation of annual percentage changes (APC) and average annual percentage changes (AAPC) in different segments, which can reveal whether a trend is increasing, decreasing, or stable. Statistical analyses were conducted using Jointpoint software (4.9.1.0) [19]. A *p*-value of < 0.05 was considered statistically significant.

### 2.3 Decomposition analysis

In public health research, decomposition analysis is commonly used to study the impact of population changes on health indicators, such as disease incidence and mortality rates. Here, we employed the decomposition analysis method proposed by Das Gupta, combined with the improved approach introduced by Cheng et al. in 2020, to dissect changes in the incidence and mortality among the MSM population into three key determinants at the population level: population aging, population growth, and epidemiological change, to quantify the impact of each factor on the overall change [20][21].

### 2.4 Autoregressive integrated moving average (ARIMA) model

The ARIMA model is a widely used statistical tool for analyzing and forecasting time series data, effectively capturing complex patterns and trends based on past observations [22]. It combines three components: Autoregression (AR), which relates the current value to past values; Integration (I), which involves differencing to achieve stationarity; and Moving Average (MA), which models the relationship between current values and past forecast errors. Typically denoted as ARIMA (*p*, *d*, *q*), where *p* represents autoregressive terms, *d* the order of differencing, and *q* the moving average terms. The model with the lowest Akaike Information Criterion (AIC) was selected as the optimal predictive model using the “auto.arima()” function [23]. We set the forecast period to 15 years, with the primary measures being Age-Standardized Incidence Rate (ASIR) and Age-Standardized Death Rate (ASDR). The model was evaluated using the white noise test, and the residual test of *p* > 0.05 indicated that it passed the white noise test. Predictions were made using the “forecast” R package.

### 2.5 Patient and public involvement

Since the study relied on publicly available aggregate data, no patients participated in defining the research question or outcome measures, nor were they involved in the study’s design or implementation.

### 2.6 Statistical analysis

Here, it’s important to note that the rate data in the GBD database is expressed per 100,000 population. Furthermore, age-standardization is a crucial statistical method in clinical data analysis that helps researchers account for the confounding effects of age, thereby allowing for a more accurate understanding of the roles and impacts of other variables. A linear regression model was established to describe the relationship between time and the natural logarithm (ln) of the age-standardized rate: *Y = αx + β + ε*, where *Y* represents the ln (age-standardized rate), *x* is the calendar year, and *ε* is the error term. Lastly, the GBD calculates UI values by repeatedly sampling and estimating data under multiple assumptions. Statistical analyses were performed using R software (version 4.3.1), with data visualization facilitated by “ggplot2” (version 3.5.1) for graphical representation and “ggsci” (version 3.2.0) for scientific color palettes [24].

## 3. Results

### 3.1 Description of MSM incidence, prevalence, and mortality in China

In 2021, there were a total of 81,219 (95% UI: 42,975–109,989) MSM cases in China, with 5,373 (95% UI: 2,849–7,106) deaths attributed to MSM-related conditions that year (Table 1). Over the period from 1990 to 2021, three key measures among the MSM population in China showed significant changes (Figure 1,2 and Supplementary figure 1). Both incidence and prevalence rates increased substantially, with the incidence rate rising from 0.36 in 1990 to 0.68 in 2021, and the prevalence rate jumping from 0.67 to 4.16. This trend indicates a considerable increase in both diagnosis and survival rates of affected individuals. Although the number of deaths increased, the mortality rate slightly decreased from 0.31 to 0.27 (Table 1). Analyzing the percentage change in ASRs per 100,000 people from 1990 to 2021, the overall incidence rate grew by 0.89, with a higher increase observed in females (0.96) compared to males (0.84). The prevalence rate showed a notable rise of 5.18, reflecting a greater number of individuals either surviving with or being diagnosed with the disease. Despite the rise in incidence, the mortality rate decreased by 0.11, with a reduction of 0.17 in males and 0.05 in females, which may be attributable to advances in medical care (Supplementary table 1).

**Figure 1.**
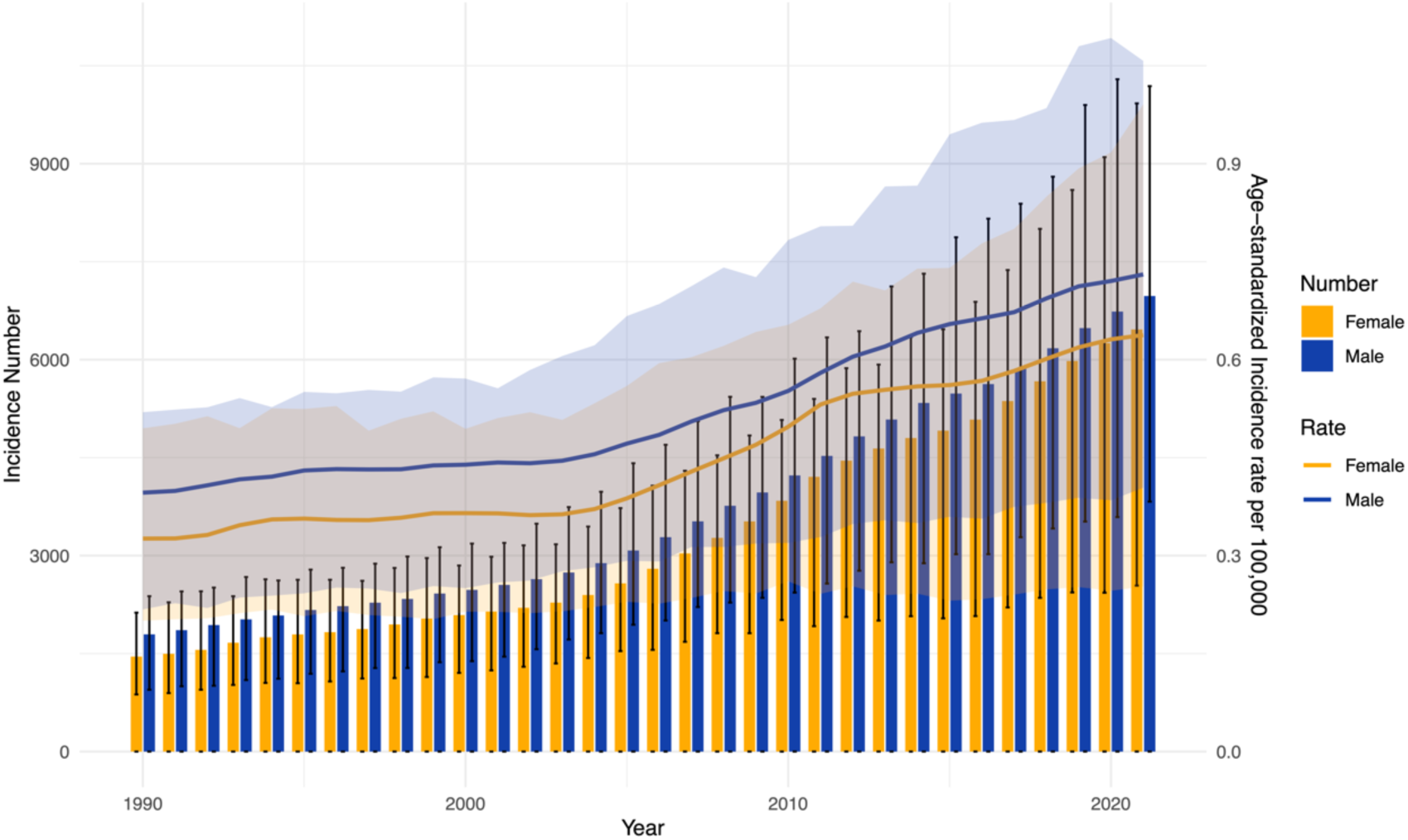
Trends in the incidence number and age-standardized incidence rate (ASIR) of MSM by gender in China, from 1990 to 2021. The bars represent the incidence number for males (blue) and females (orange), while the lines show the ASIR per 100,000 individuals. Shaded areas around the lines indicate the uncertainty intervals for ASIR estimates.

**Figure 2.**
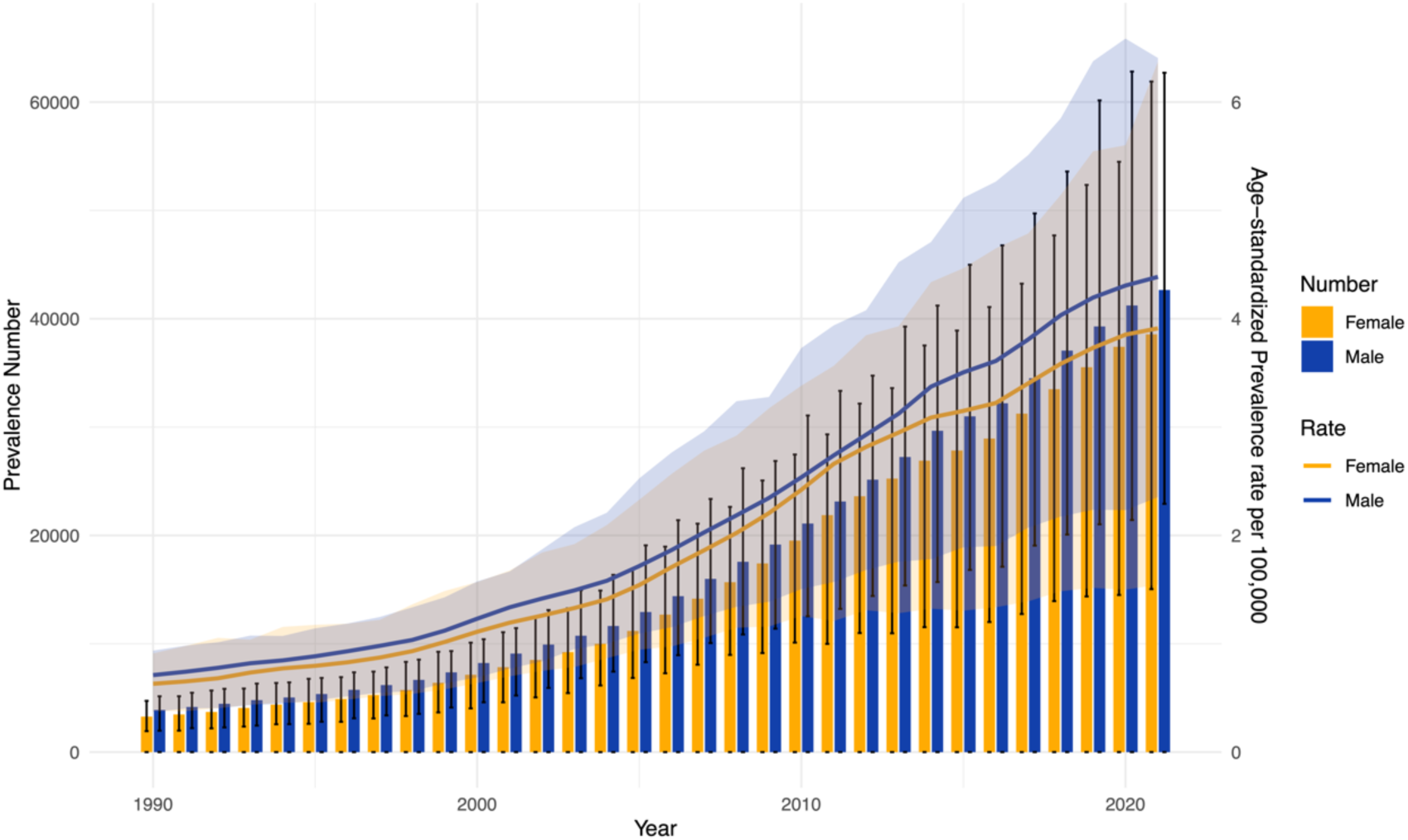
Trends in the prevalence number and age-standardized prevalence rate (ASPR) of MSM by gender in China, from 1990 to 2021. The bars represent the prevalence number for males (blue) and females (orange), while the lines show the ASPR per 100,000 individuals. Shaded areas around the lines indicate the uncertainty intervals for ASPR estimates.

**Table 1.**
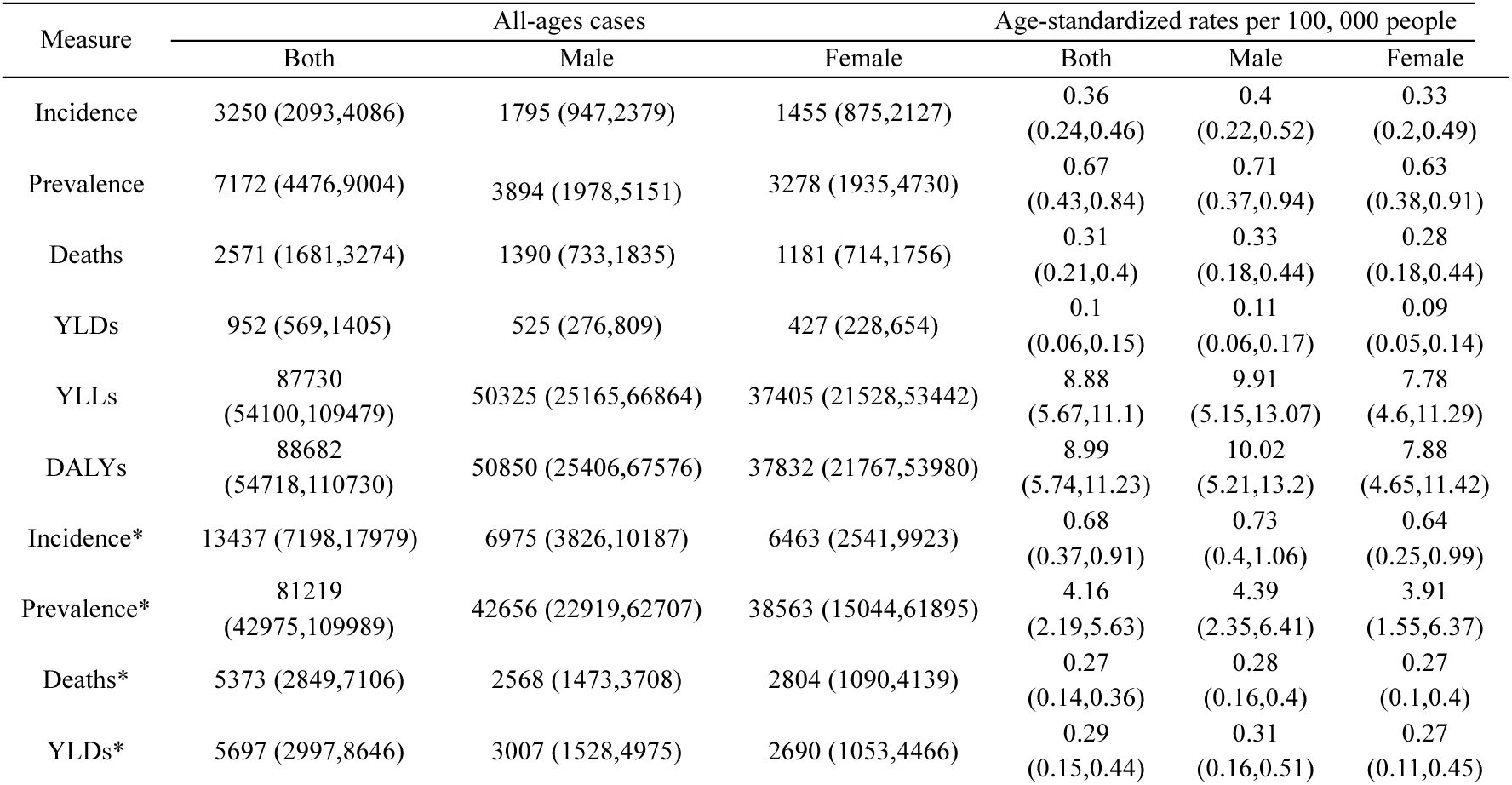
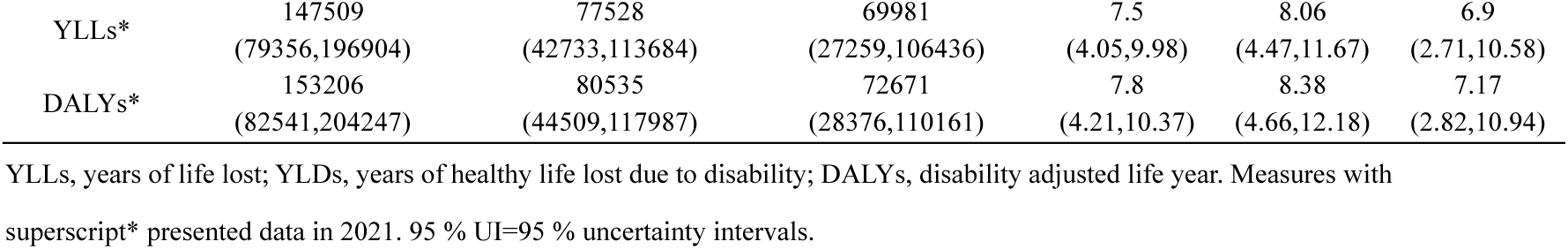
Incidence, prevalence, deaths, YLDs, YLLs and DALYs for malignant skin melanoma in 1990 and 2021 (all-ages and age-standardized rates data was included)

| Measure | All-ages cases |  |  | Age-standardized rates per 100, 000 people |  |  |
| --- | --- | --- | --- | --- | --- | --- |
|  | Both | Male | Female | Both | Male | Female |
| Incidence | 3250 (2093,4086) | 1795 (947,2379) | 1455 (875,2127) | 0.36<br>(0.24,0.46) | 0.4<br>(0.22,0.52) | 0.33<br>(0.2,0.49) |
| Prevalence | 7172 (4476,9004) | 3894 (1978,5151) | 3278 (1935,4730) | 0.67<br>(0.43,0.84) | 0.71<br>(0.37,0.94) | 0.63<br>(0.38,0.91) |
| Deaths | 2571 (1681,3274) | 1390 (733,1835) | 1181 (714,1756) | 0.31<br>(0.21,0.4) | 0.33<br>(0.18,0.44) | 0.28<br>(0.18,0.44) |
| YLDs | 952 (569,1405) | 525 (276,809) | 427 (228,654) | 0.1<br>(0.06,0.15) | 0.11<br>(0.06,0.17) | 0.09<br>(0.05,0.14) |
| YLLs | 87730<br>(54100,109479) | 50325 (25165,66864) | 37405 (21528,53442) | 8.88<br>(5.67,11.1) | 9.91<br>(5.15,13.07) | 7.78<br>(4.6,11.29) |
| DALYs | 88682<br>(54718,110730) | 50850 (25406,67576) | 37832 (21767,53980) | 8.99<br>(5.74,11.23) | 10.02<br>(5.21,13.2) | 7.88<br>(4.65,11.42) |
| Incidence* | 13437 (7198,17979) | 6975 (3826,10187) | 6463 (2541,9923) | 0.68<br>(0.37,0.91) | 0.73<br>(0.4,1.06) | 0.64<br>(0.25,0.99) |
| Prevalence* | 81219<br>(42975,109989) | 42656 (22919,62707) | 38563 (15044,61895) | 4.16<br>(2.19,5.63) | 4.39<br>(2.35,6.41) | 3.91<br>(1.55,6.37) |
| Deaths* | 5373 (2849,7106) | 2568 (1473,3708) | 2804 (1090,4139) | 0.27<br>(0.14,0.36) | 0.28<br>(0.16,0.4) | 0.27<br>(0.1,0.4) |
| YLDs* | 5697 (2997,8646) | 3007 (1528,4975) | 2690 (1053,4466) | 0.29<br>(0.15,0.44) | 0.31<br>(0.16,0.51) | 0.27<br>(0.11,0.45) |
| YLLs* | 147509<br>(79356,196904) | 77528<br>(42733,113684) | 69981<br>(27259,106436) | 7.5<br>(4.05,9.98) | 8.06<br>(4.47,11.67) | 6.9<br>(2.71,10.58) |
| DALYs* | 153206<br>(82541,204247) | 80535<br>(44509,117987) | 72671<br>(28376,110161) | 7.8<br>(4.21,10.37) | 8.38<br>(4.66,12.18) | 7.17<br>(2.82,10.94) |

YLLs, years of life lost; YLDs, years of healthy life lost due to disability; DALYs, disability adjusted life year. Measures with superscript\* presented data in 2021. 95 % UI=95 % uncertainty intervals.

Additionally, the YLDs (Years Lived with Disability) and YLLs (Years of Life Lost) rose significantly over this period, leading to an increase in DALYs (Disability-Adjusted Life Years) from 88,682 in 1990 to 153,206 in 2021. This indicates that the health burden of MSM on Chinese society has intensified over the years (Table 1).

From a gender perspective, incidence and prevalence rates in males consistently surpassed those in females. In 1990, the incidence rate was 0.4 in males and 0.33 in females; by 2021, these rates had risen to 0.73 for males and 0.64 for females. Although mortality rates showed a slight downward trend for both genders, the mortality rate among males (declining from 0.33 to 0.28) remained higher than that of females (decreasing from 0.28 to 0.27). Furthermore, the DALYs and YLLs among males were also higher than among females, suggesting that males not only face a higher risk of disease but also endure a more significant health burden from MSM-related conditions (Table 1). Subsequent analysis divided patients by age group in 1990 and 2021 to identify age-specific high-risk groups for incidence, prevalence, and mortality across genders. In 1990, the distribution was relatively balanced between males and females, with a slightly higher incidence among males in the 40–44 (male: 220; female: 111) and 55–59 (male: 284; female: 171) age groups, predominantly affecting middle-aged individuals (Figure 3A). By 2021, the incidence of new cases showed a marked increase compared to 1990, especially in older age groups (Figure 3C). The age group with the highest incidence centered around 55–59 (male: 1,095; female: 774), with negligible risk below the 25–29 age group. A similar trend was observed in prevalence, with the highest rate occurring in the 55–59 age group (male: 6,856; female: 5,322) in 2021 (Figure 3D). Regarding mortality, in 2021, the highest numbers of deaths were recorded among males aged 55–59 (394) and females aged 75–79 (362) (Supplementary figure 3,4). This analysis underscores the growing health burden among MSM populations in China, with notable differences between genders and a shift toward older age groups over time (Supplementary figure 5,6).

**Figure 3.**
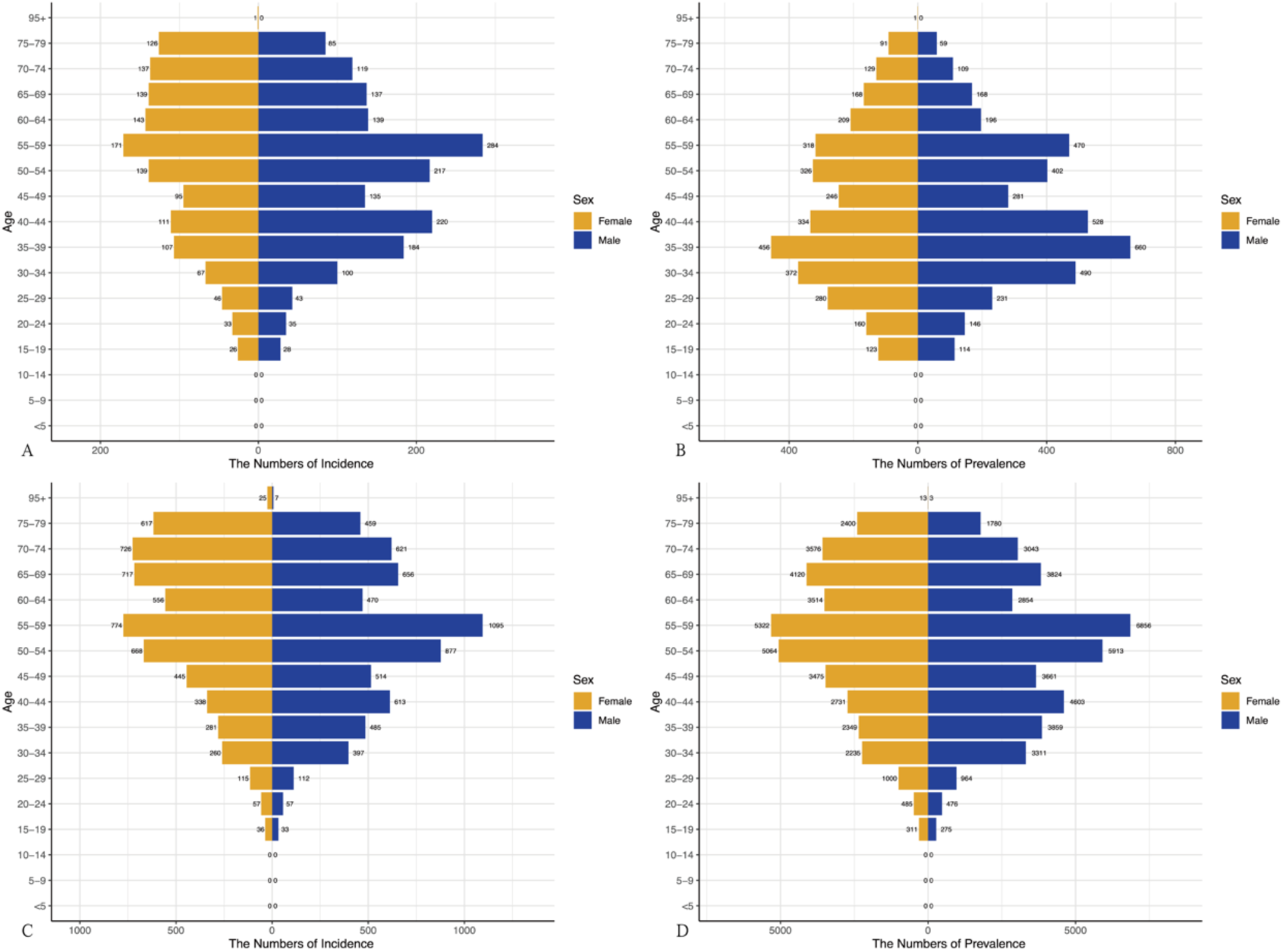
Distribution of disease burden among MSM by age group and sex in 1990 and 2021. A. Incidence of new cases among MSM by age group and sex in 1990. B. Prevalence of cases among MSM by age group and sex in 1990. C. Incidence of new cases among MSM by age group and sex in 2021. D. Prevalence of cases among MSM by age group and sex in 2021.

To gain a deeper understanding of incidence and mortality trends, we segmented the time period from 1991 to 2021. Initially, prevalence rates exhibited a relatively stable trend in 1991, but over time, risk levels across different age groups became more distinct. From 1991 to 2021, prevalence rates increased with age, peaking within the 50-54 age group, then gradually declining in older groups (Figure 4A, C). Mortality counts reached their maximum within the 55-64 age group, subsequently decreasing as age increased further. Over time, prevalence rates and mortality counts increased across all age groups, with a notable acceleration post-2001, indicating a worsening health trend within this population (Figure 4B, D). Additionally, individuals born after 1905 experienced significant increases in prevalence across age groups (Figure 5A). For those born between 1930 and 1970, mortality counts substantially increased in the 50-74 age range, particularly among middle-aged individuals, where mortality peaked. In contrast, cohorts born after 1980 showed markedly lower mortality across all age groups, suggesting a potential improvement in health outcomes, possibly due to advancements in healthcare or preventive measures (Figure 5B).

**Figure 4.**
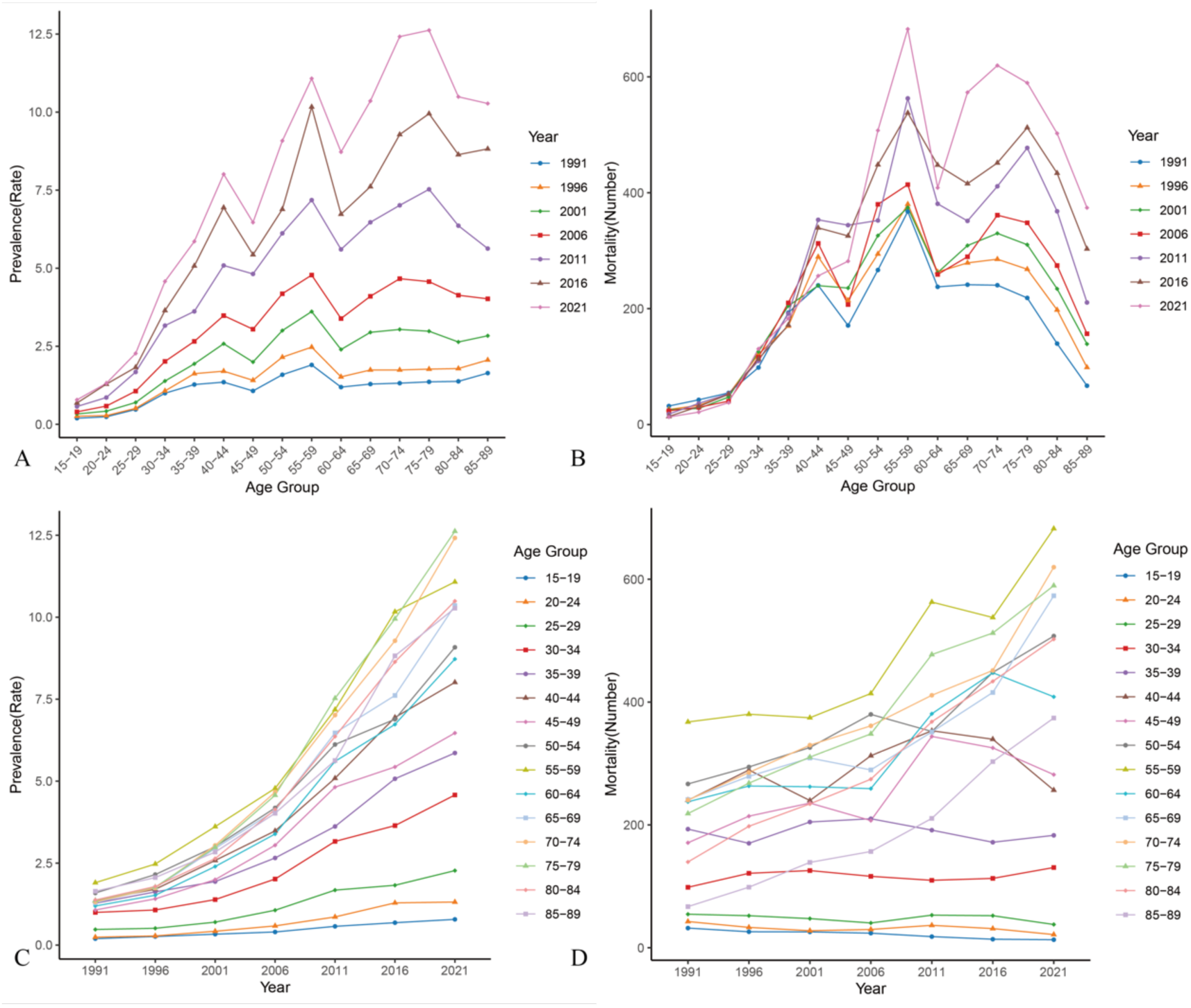
Trends in prevalence rates and mortality number among MSM by age group over time. A. Prevalence rates (per 100,000) of MSM across different age groups from 1991 to 2021. B. Mortality number of MSM across different age groups from 1991 to 2021. C. Prevalence rates (per 100,000) among MSM by year, segmented by age group from 15-19 to 85-89. D. Mortality number among MSM by year, with age groups segmented similarly.

**Figure 5.**
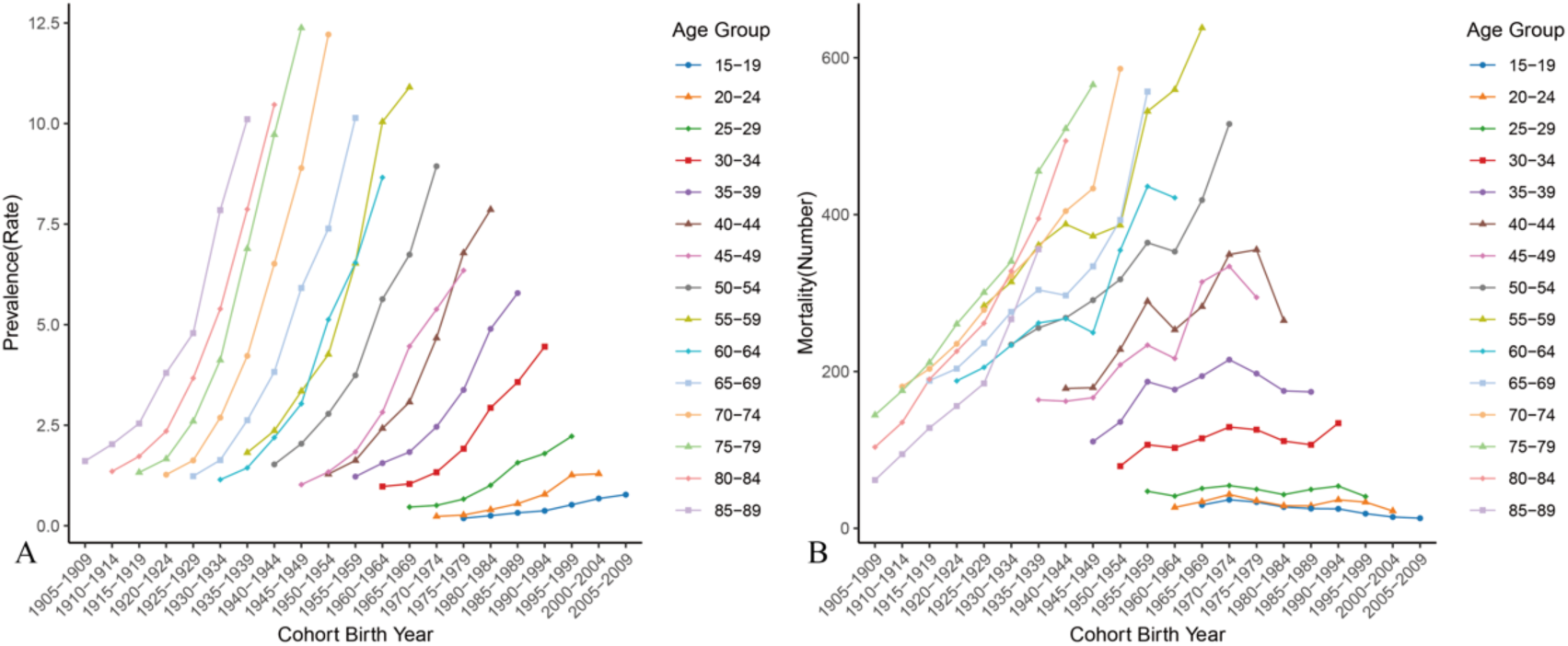
Trends in cohort-based MSM prevalence rates and mortality number among MSM by age group over time. Back-calculating birth years based on obtained age intervals. A. Prevalence rates (per 100,000) of MSM across different birth time from 1905 to 2009. B. Mortality number of MSM across different birth time from 1905 to 2009.

### 3.2 Joinpoint regression analysis and decomposition analysis of MSM incidence and mortality

From 1990 to 2021, the age-standardized incidence rate of malignant melanoma in China exhibited an overall upward trend. Between 1990 and 1994, the average annual growth rate of incidence was 2.06%, which then decelerated to 0.47% from 1994 to 2004. However, starting in 2004, the rate of increase significantly accelerated, reaching an average annual growth rate of 4.39% between 2004 and 2012, marking the period of fastest growth. After 2012, the growth rate slightly slowed to 1.98% per year, but the incidence continued to rise (Figure 5A). Similarly, the age-standardized mortality rate for malignant melanoma also showed an overall increasing trend from 1990 to 2021. Initially, there was moderate growth from 1990 to 1998, with an APC of 4.87%, followed by a sharp increase to 9.12% from 1998 to 2001—the highest growth observed. The APC then decelerated to 5.67% from 2001 to 2004, accelerated again to 8.82% from 2004 to 2011, and gradually slowed to 5.91% between 2011 and 2014, reaching a low of 3.97% from 2014 to 2021 (Figure 5B). The two upward trends may be closely associated with factors such as population aging, environmental changes, and advancements in diagnostic technologies, indicating a growing burden of malignant melanoma in China. All statistical analyses are significant.

Decomposition analysis suggests that the increases in both incidence and mortality rates are primarily driven by population aging, with a more pronounced effect on males (Figure 6). While epidemiological shifts contribute to the rise in incidence and mortality, they play a smaller role compared to aging (Figure 7A, B). The impact of population growth is minimal, underscoring aging as a critical factor driving the increasing disease burden.

**Figure 6.**
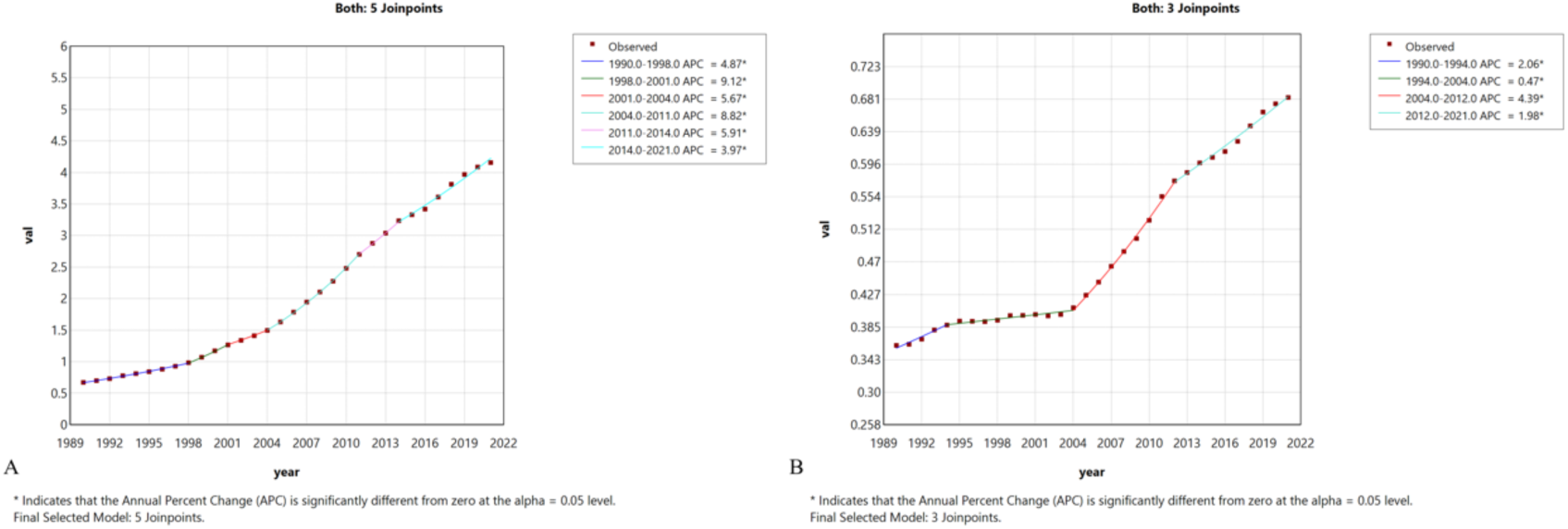
Joinpoint regression analysis of prevalence and incidence trends for MSM from 1990 to 2021. A. Incidence of MSM over time, with 3 joinpoints identified. B. Prevalence of MSM over time, with 5 joinpoints identified. All Annual Percent Change (APC) values with a superscript* are statistically significant.

**Figure 7.**
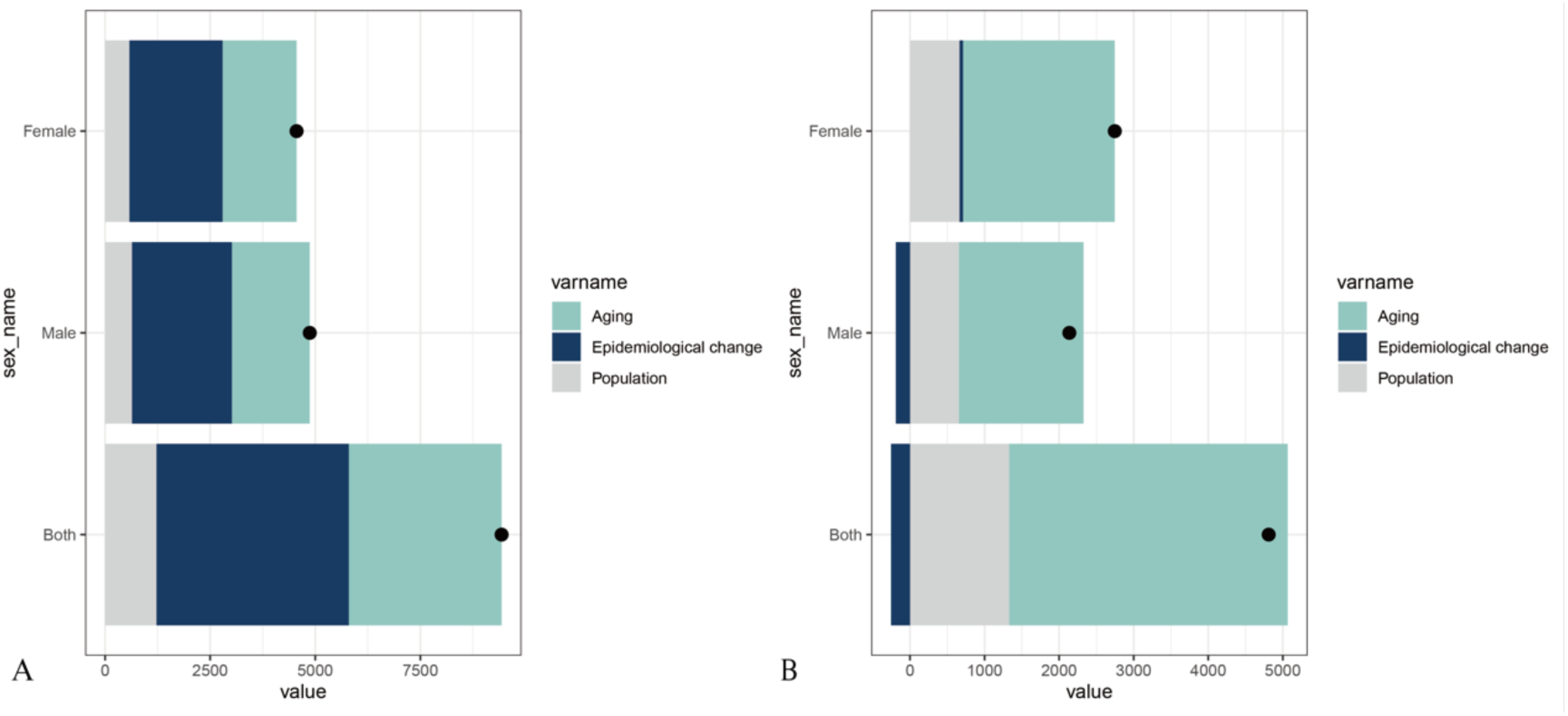
A. Decomposition analysis of the impact of aging, epidemiological change, and population growth on MSM-related incidence and mortality stratified by sex. B. Decomposition analysis of the impact of aging, epidemiological change, and population growth on MSM-related incidence and mortality stratified by sex. Each bar is segmented by color to indicate the relative contribution of each factor: aging (dark blue), epidemiological change (teal), and population growth (gray). Black dots represent the total values for each subgroup.

### 3.3 ARIMA analysis of MSM ASIR and ASDR

From 1990 to 2036, based on observed data from 1990 to 2021, both male and female ASIR trends indicate a steady increase. Projections suggest that by 2036, the ASIR will reach 0.88 for males and 0.71 for females (Figure 8A, B). For ASDR, historical data shows stability for both sexes from 1990 to 2021, with forecasts indicating that by 2036, ASDR will decline to 0.25 for males and 0.26 for females (Figure 8C, D) (Supplementary table 2,3). The forecast confidence intervals (yellow shaded areas) highlight some degree of uncertainty in these projections, particularly notable in the ASIR forecasts.

**Figure 8.**
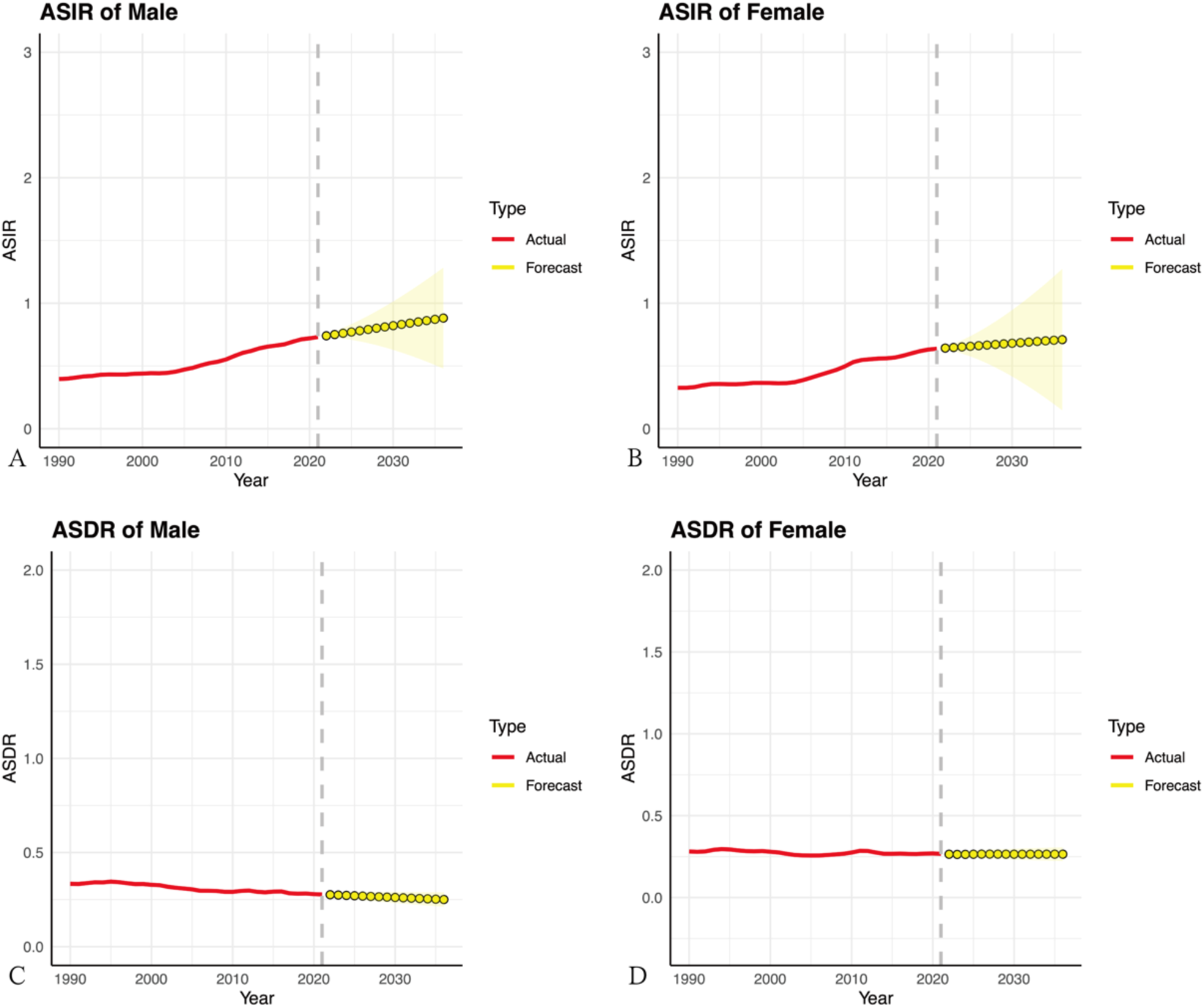
ARIMA model forecasts of Age-Standardized Incidence Rate (ASIR) and Age-Standardized Death Rate (ASDR) for MSM by sex, 1990–2036. A. Forecast of the ASIR for males from 1990 to 2036. B. Forecast of the ASIR for females from 1990 to 2036. C. Forecast of the ASDR for males from 1990 to 2036. D. Forecast of the ASDR for females from 1990 to 2036. The red line represents actual ASIR data up to 2021, while the yellow shaded area indicates forecasted values with a confidence interval from 2022 to 2036.

## 4. Discussion

In 2021, China reported an estimated 13,437 new cases of MSM (95% UI: 7,198– 17,979), with a total MSM patient population of 81,219 (95% UI: 42,975–109,989). That same year, MSM-related deaths numbered 5,373 (95% UI: 2,849–7,106). From 1990 to 2021, all three key measures among Chinese MSM exhibited significant changes: incidence and prevalence rates increased substantially, while mortality rates declined slightly DALYs rose from 88,682 in 1990 to 153,206 in 2021, indicating an increasing health burden of MSM on Chinese society. Moreover, incidence, prevalence, and mortality rates for males consistently exceeded those for females, highlighting a higher susceptibility to disease and a greater health risk from malignant melanoma among males. The highest prevalence rate in China was observed in the 55-59 age group (male: 6,856; female: 5,322). Regarding mortality, the highest numbers in 2021 were among males aged 55-59 (394 deaths) and females aged 75-79 (362 deaths). Joinpoint regression analysis identified 2004-2012 as the period of the most rapid ASIR increase, with an annual percentage change (APC) of 4.39%, while the fastest rise in ASDR occurred from 2004 to 2011, with an APC of 8.82%. Decomposition analysis suggests that the growth in incidence and mortality rates of malignant melanoma in China is primarily driven by an aging population, with a notably stronger impact on males. ARIMA projections indicate that by 2036, the ASIR will reach 0.88 for males and 0.71 for females. Meanwhile, historical data from 1990 to 2021 shows stable ASDR rates for both sexes, with projections suggesting a decline to 0.25 for males and 0.26 for females by 2036. Overall, despite advancements in treatment and survival, the health burden of MSM in China is likely to continue increasing due to population aging. Males, particularly middle-aged men, face a higher risk of developing the disease. This calls for targeted interventions to reduce MSM incidence and mortality rates in this vulnerable group.

In this study, we observed that the incidence of MSM in China has been on the rise in recent years, while mortality rates have shown a gradual decline. We attribute this mainly to advancements in medical technology, which have made early diagnosis more accessible, as well as to innovations in immunotherapy and targeted therapy. Melanoma and nevi have similar clinical presentations, making it challenging for dermatologists to differentiate between them, which entails the use of advanced diagnostic techniques [25]. In China, dermoscopy has been widely adopted, and domestic researchers have made significant progress in this field. Dermoscopy enhances the accuracy of diagnosing tumor-related diseases, evaluates the effectiveness of treatments, and determines treatment endpoints [26]. It also aids in the differential diagnosis of inflammatory diseases and assesses disease severity [27]. By incorporating dermoscopy into clinical examinations, the sensitivity of clinical melanoma diagnosis has improved from around 60% to 90% [28]. Furthermore, the Chinese Skin Image Database, launched in 2017 as a working platform for dermatologists, has promoted the development of dermoscopy in China [29][30].

In 2018, researchers employed cluster sampling to recruit first-year students from five universities across various regions of China to assess Chinese adolescents’ knowledge of melanoma. A total of 21,086 questionnaires were completed by university students, with an average age of 18.0 ± 0.8 years. The health literacy and attitudes mean scores were 9.83 ± 7.46 (maximum score: 28) and 16.98 ± 2.92 (maximum score: 20), respectively. This indicates that health literacy related to melanoma among Chinese university students is generally insufficient and needs improvement. Recognizing this early on, Chinese dermatologists have worked towards increasing awareness, and over the years, melanoma education and outreach efforts have gradually matured [31]. A growing number of face-to-face and online training courses enhancing dermatologists’ diagnostic skills have emerged [30].

Although melanoma is considered a rare disease in China, with a lower incidence compared to Caucasian populations, significant subtype differences between Chinese and American melanoma contribute to a higher malignancy and mortality rate in China compared to the West [31]. Approximately 25% of advanced melanoma patients in China carry BRAF gene mutations, which are associated with higher malignancy and often poorer clinical outcomes [32]. This clear mutation target has opened a new era of targeted therapy with BRAF inhibitors in improving treatment efficacy [33]. Thanks to the strong support from China’s healthcare policies for innovative melanoma drugs, advanced therapies like dabrafenib combined with trametinib allow Chinese melanoma patients to receive treatment on par with international standards under insurance coverage. At the 2020 European Society for Medical Oncology annual meeting, updated data on dabrafenib combined with trametinib for treating advanced Asian patients with BRAF V600 mutant melanoma showed an objective response rate of 71.7%, a median progression-free survival of 9.3 months, and a median overall survival of 21.1 months, underscoring the effectiveness of dual-targeted therapy in Chinese patients with advanced melanoma [32]. The balanced distribution of medical resources allows more patients to receive timely treatment. Coupled with enhanced health education and increased prevention awareness, high-risk populations can adopt effective protective measures. Additionally, new options for patient treatment have provided new treatment options for patients, thus improving overall survival rates.

Previous research has indicated that men are approximately 1.5 times more likely than women to develop melanoma and have poorer prognoses. Eirini Chrysanthou et al. collected and analyzed data from seven online datasets, covering normal skin, common acquired nevi, and melanoma. Clear differences were observed between sexes and stages regarding survival-associated genes, transcriptomic profiles, and variability, underscoring the sexual dimorphism in gene expression [34]. In a cross-sectional study using data from eight predominantly fair-skinned populations, it was found that melanoma incidence rates among women were higher than men before middle age in all countries. This finding contrasts sharply with ours, which indicates that, in China, the incidence rate among women is lower than that of men across all age groups. Currently, no studies support this evidence [35]. Melanoma on the male trunk and female lower limbs is likely genetically determined and less related to sun exposure. This suggests that melanoma incidence among Chinese patients is primarily driven by genetics [36]. Therefore, identifying sex-differentiated genes in Chinese melanoma patients is particularly important.

At the recent CSCO conference, new treatment strategies were proposed to address the growing number of melanoma patients. For advanced melanoma patients, treatment strategies should focus on two primary aspects. First, for various groups such as those with BRAF or KRAS mutations, it is essential to further explore and optimize combination therapies based on current regimens to enhance patient benefits [37]. Additionally, it is crucial to accelerate drug development by targeting novel sites identified in prior basic research, providing patients with more therapeutic options. Future drug development could enhance efficacy and reduce toxicity through structural modifications or formulation optimizations [38]. Targeted therapies are known for their precision; however, drug resistance remains a challenge. The trend towards combining targeted therapies with immunotherapy for melanoma treatment is well established [39]. There are two main approaches: delaying resistance as much as possible in frontline treatment, and when resistance arises, seeking effective therapeutic strategies by rationally utilizing existing methods or replacing frontline treatments with alternative approaches. In frontline treatment, a strategy involving dual-target inhibitors combined with immunotherapy can be employed to delay resistance. Clinical practice has shown that this combined approach can delay the onset of resistance and extend overall survival [40][41]. For second-line treatment after resistance occurs, it is recommended to perform rebiopsy and molecular testing to determine whether the resistance mechanism involves MAPK pathway reactivation or an alternative pathway [42]. The appropriate treatment approach can then be selected based on the specific resistance mechanism. Here, we recommend the government take the following actions: increase funding for fundamental and clinical research on skin cancer, with a particular focus on emerging therapies such as immunotherapy and targeted treatments. Support genetic research for high-risk patients to better understand the underlying causes and advance precision medicine. Improve the skin cancer registry system to enhance data monitoring of MSM incidence and mortality rates. Systematic data collection will enable the government and researchers to assess trends more accurately and promptly adjust prevention and control measures. It is also advisable to conduct long-term follow-up for diagnosed patients to monitor treatment outcomes and recurrence rates, thereby optimizing long-term health management strategies for patients.

This study has some limitations. First, our research did not delve into regional variations down to the provincial level within China. Secondly, although Global Burden of Disease data has been extensively standardized, uncertainties remain in data quality control—these include data collection methods, data sources, processing methods, coding, healthcare accessibility, cultural differences, and socioeconomic disparities. Such factors may affect the accuracy and robustness of disease burden assessments. Lastly, the ARIMA model predicts trends by analyzing time series, essentially forecasting linear patterns, and is therefore unable to predict sequences with turning points.

## 5. Conclusion

The burden of MSM in China has increased over the past three decades, with a rise in both incidence and prevalence rates, especially among males and older age groups. Although mortality rates have shown a modest decline, the disease remains a significant public health concern. The study underscores the need for more research on Chinese melanoma patients, as current treatments and data are often based on studies of Caucasian populations. Advancements in early detection, targeted therapies, and combination therapies with immunotherapy have shown promise in improving patient outcomes. However, drug resistance presents a major challenge. Future efforts should focus on optimizing combination therapies, expediting the development of new drugs targeting specific genetic mutations, and enhancing public awareness to prevent and manage melanoma effectively. Continued research into the genetic factors influencing melanoma in the Chinese population is essential to develop more personalized and effective treatment strategies.

## Data Availability

The Global Burden of Disease study 2021 is an open-access resource; data are available at http://ghdx.healthdata.org/gbd-results-tool.

## Abbreviations

MSM: Malignant Skin Melanoma
GBD: Global Burden of Disease
HIV: Human Immunodeficiency Viruses
CSCO: Chinese Society of Clinical Oncology
ICD-10: International Classification of Diseases 10th revision
CODEm: Cause of Death Ensemble Model
YLLs: Years of Life Lost
YLDs: Years Lived with Disability
DALYs: Disability-Adjusted Life Years
UIs: Uncertainty Intervals
APC: Annual Percentage Changes
AAPC: Average Annual Percentage Changes
ARIMA: Autoregressive Integrated Moving Average
ASIR: Age-Standardized Incidence Rate
ASDR: Age-Standardized Death Rate
Ln: Natural Logarithm

## Declaration of interests

The authors declare that they have no known competing financial interests or personal relationships that could have appeared to influence the work reported in this paper.

## Author Contribution Statement

Chenyu Zhao and Keqiang Ma contributed equally to this work. Chenyu Zhao conceptualized and drafted the manuscript. Keqiang Ma plotted the figures. Zhaoqi Wu, Ruihan Zhang, Yiting Wang and Yiting He collected the information. Martin Gluchman was responsible for language check. Hui Wang revised the manuscript. All authors read and approved the final manuscript.

## Acknowledgements

This work was supported by grants from the National Natural Science Foundation of China (No. 81702611 to Hui Wang). We appreciate the work of the Global Burden of Disease study 2021 collaborators.

## Ethical Statement

No patients were involved in defining the research question, determining outcome measures, or participating in the study’s design or implementation. Since this study utilized publicly available data, no ethical approval was required.

## Funding Information

This work was supported by grants from the National Natural Science Foundation of China (No. 81702611 to Hui Wang).

## Supplementary Materials

**Supplementary table 1.**
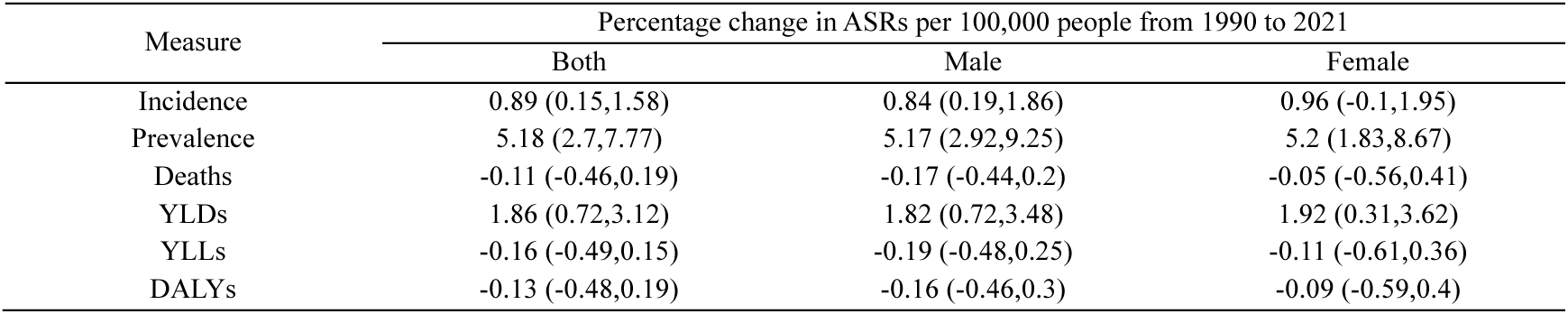
Incidence, prevalence, deaths, YLDs, YLLs and DALYs percentage change for malignant skin melanoma from 1990 to 2021. ASRs, age-standardized rates; YLLs, years of life lost; YLDs, years of healthy life lost due to disability; DALYs, disability adjusted life year. 95 % UI=95 % uncertainty intervals.

**Supplementary table 2.**
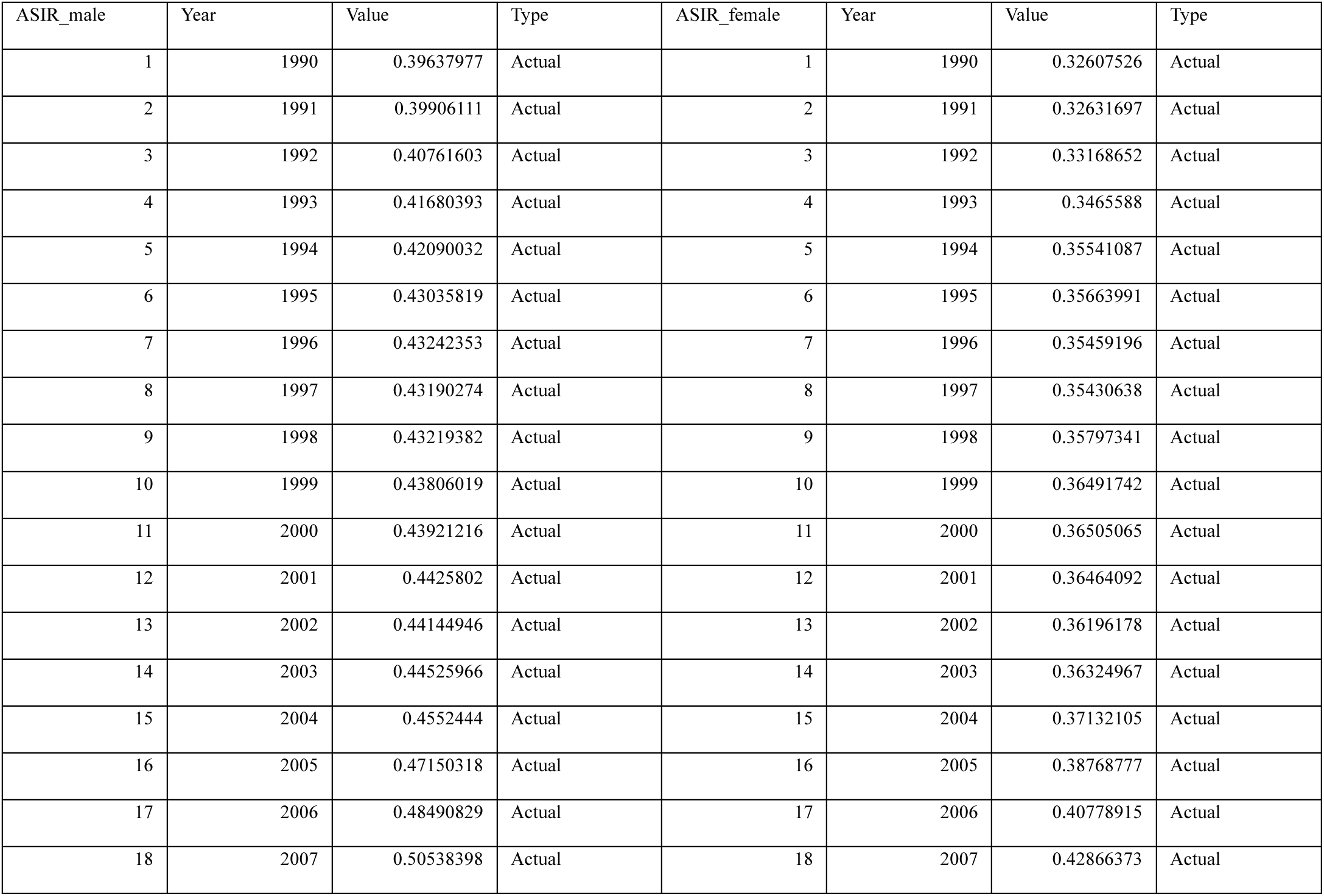
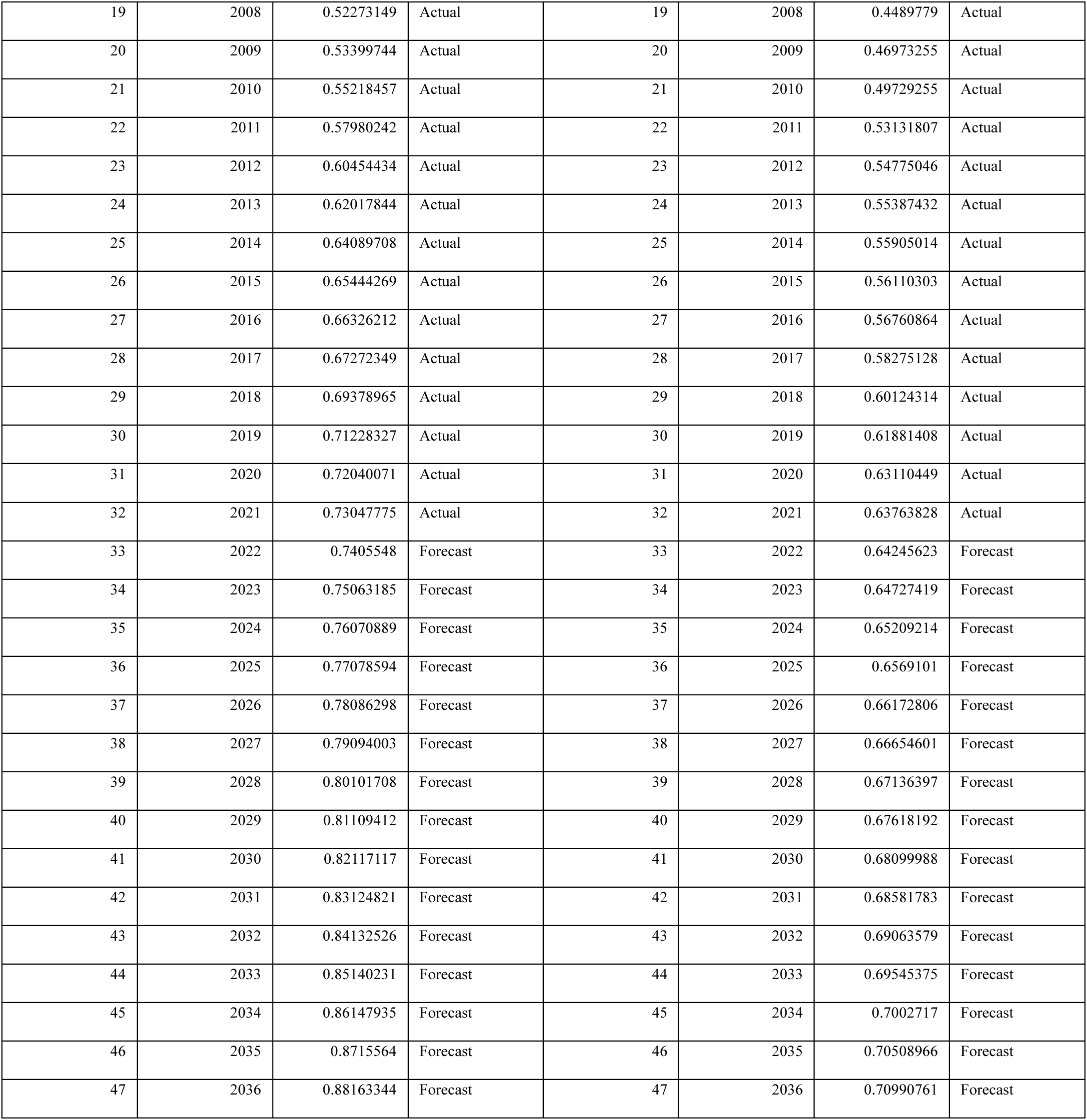
ARIMA forecast data of ASIR by gender.

**Supplementary table 3.**
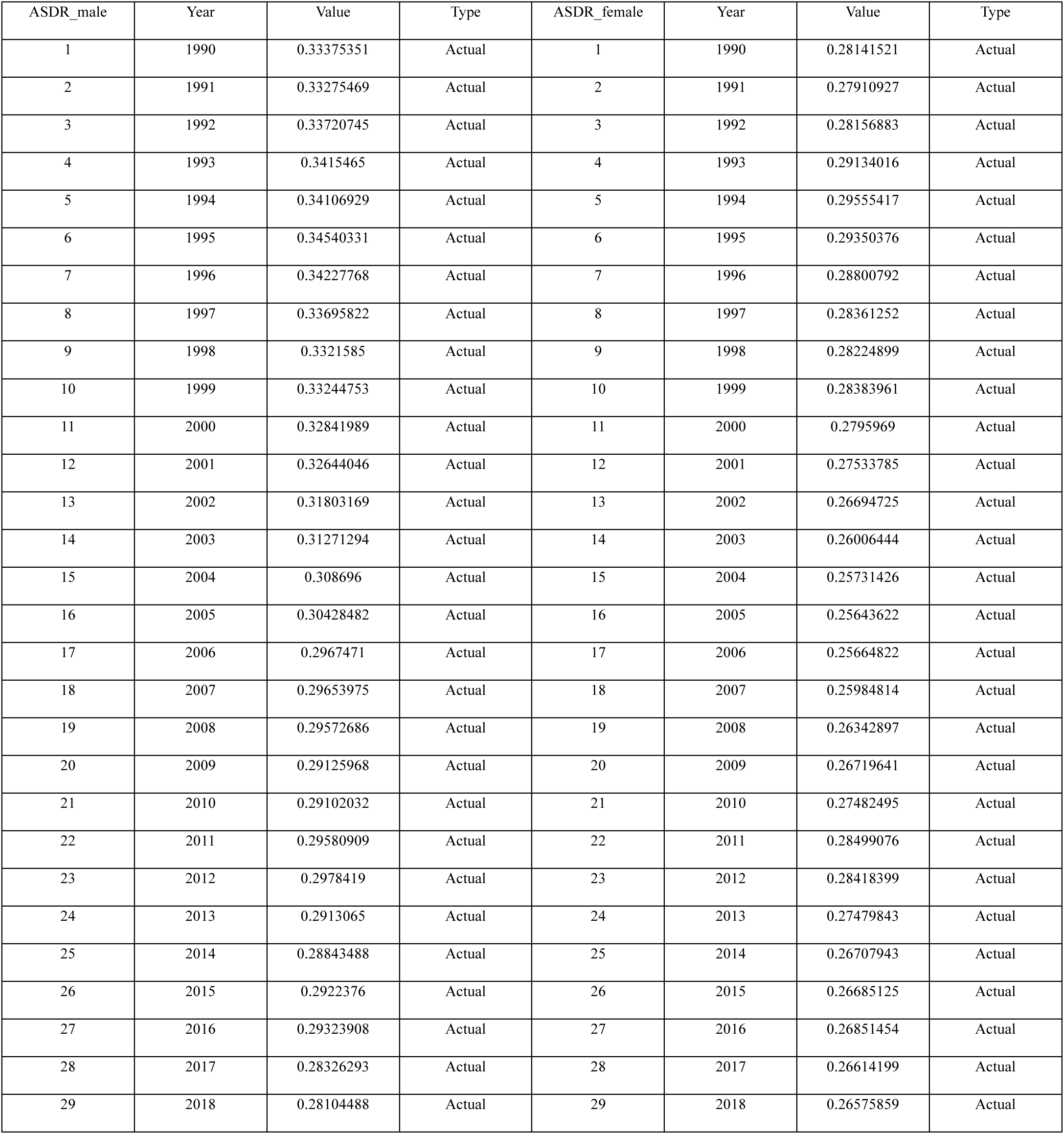
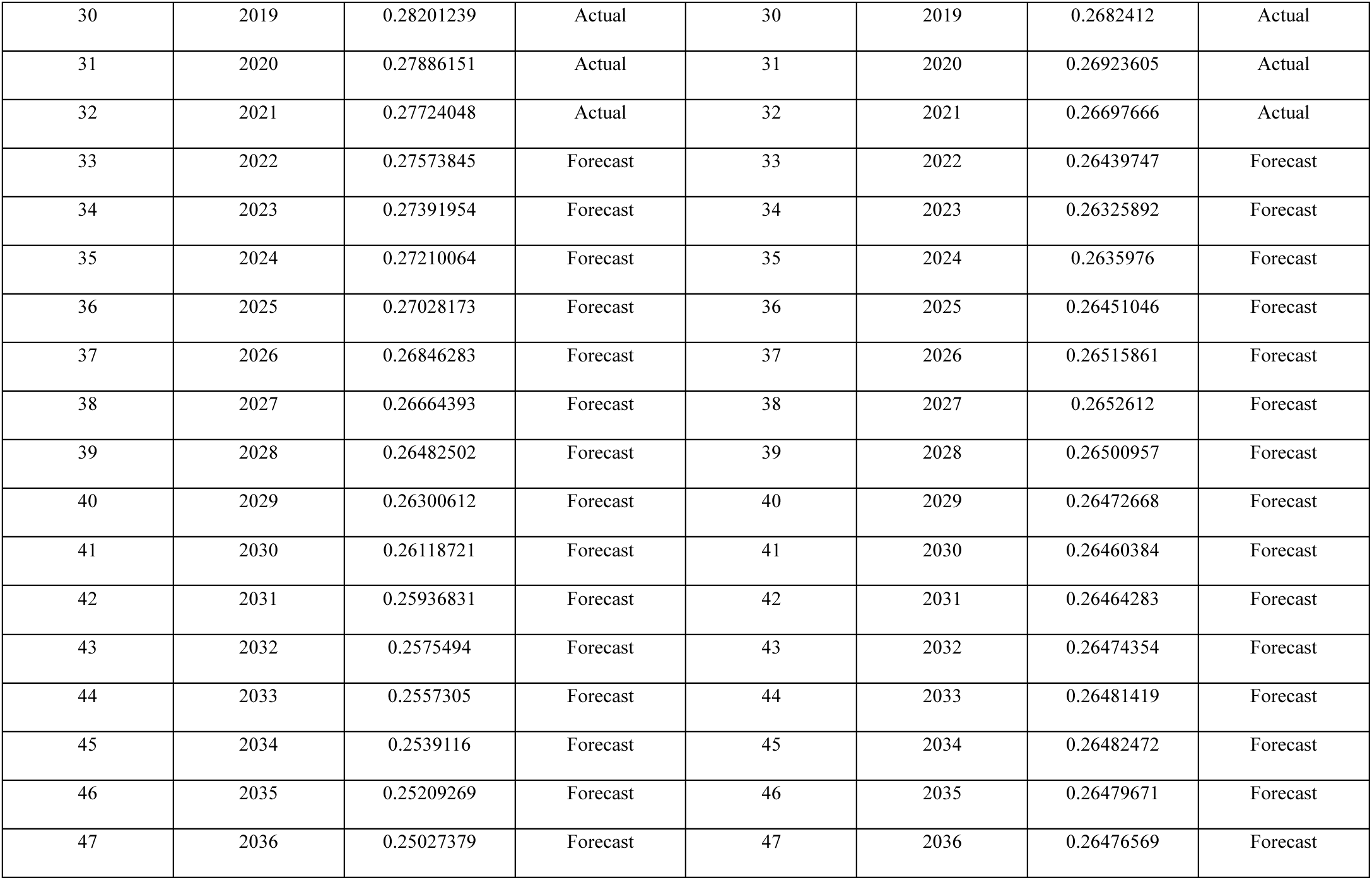
ARIMA forecast data of ASDR by gender.

**Supplementary figure 1.**
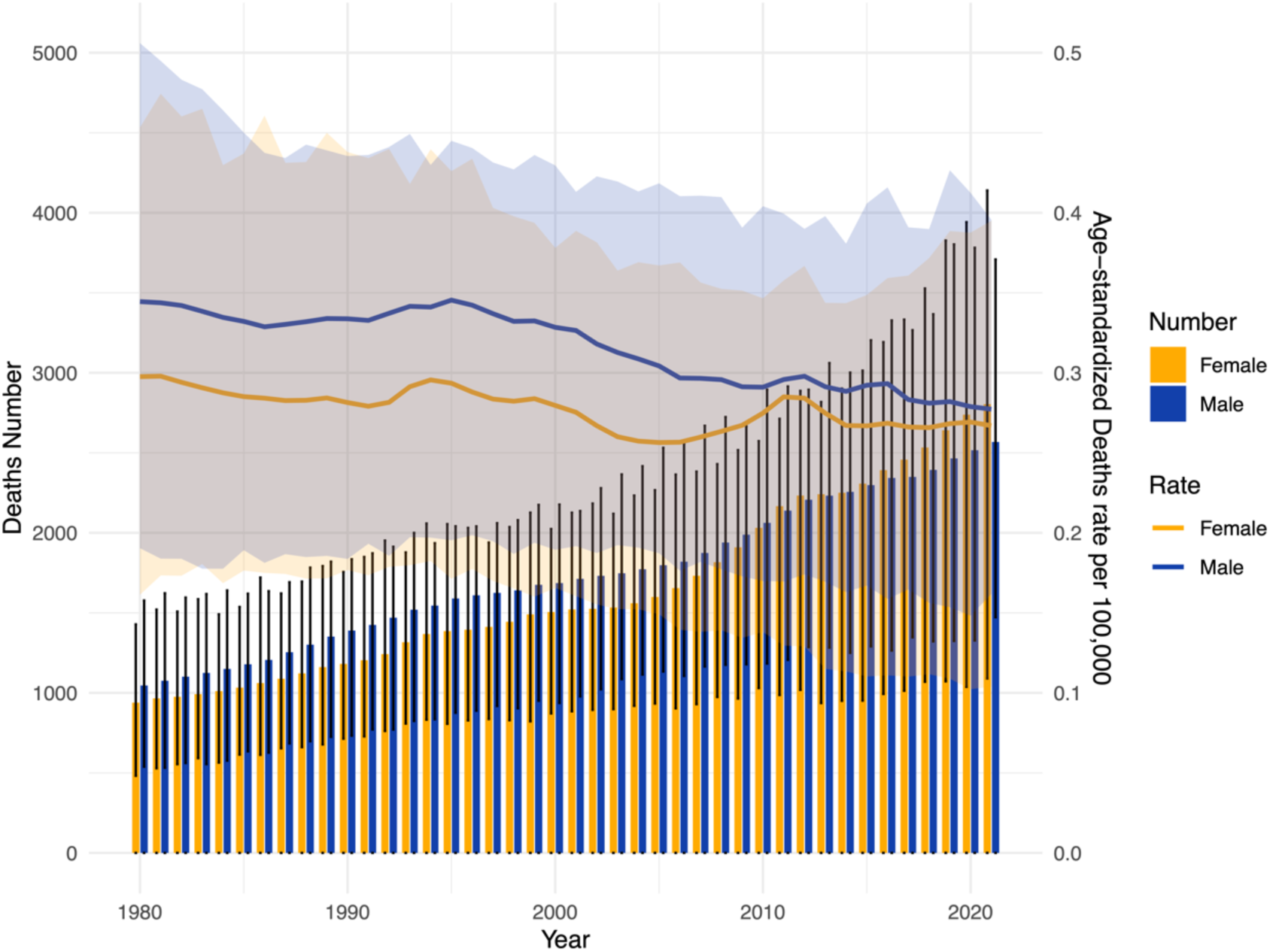
Trends in the deaths number and age-standardized deaths rate (ASDR) of MSM by gender in China, from 1990 to 2021.

**Supplementary figure 2.**
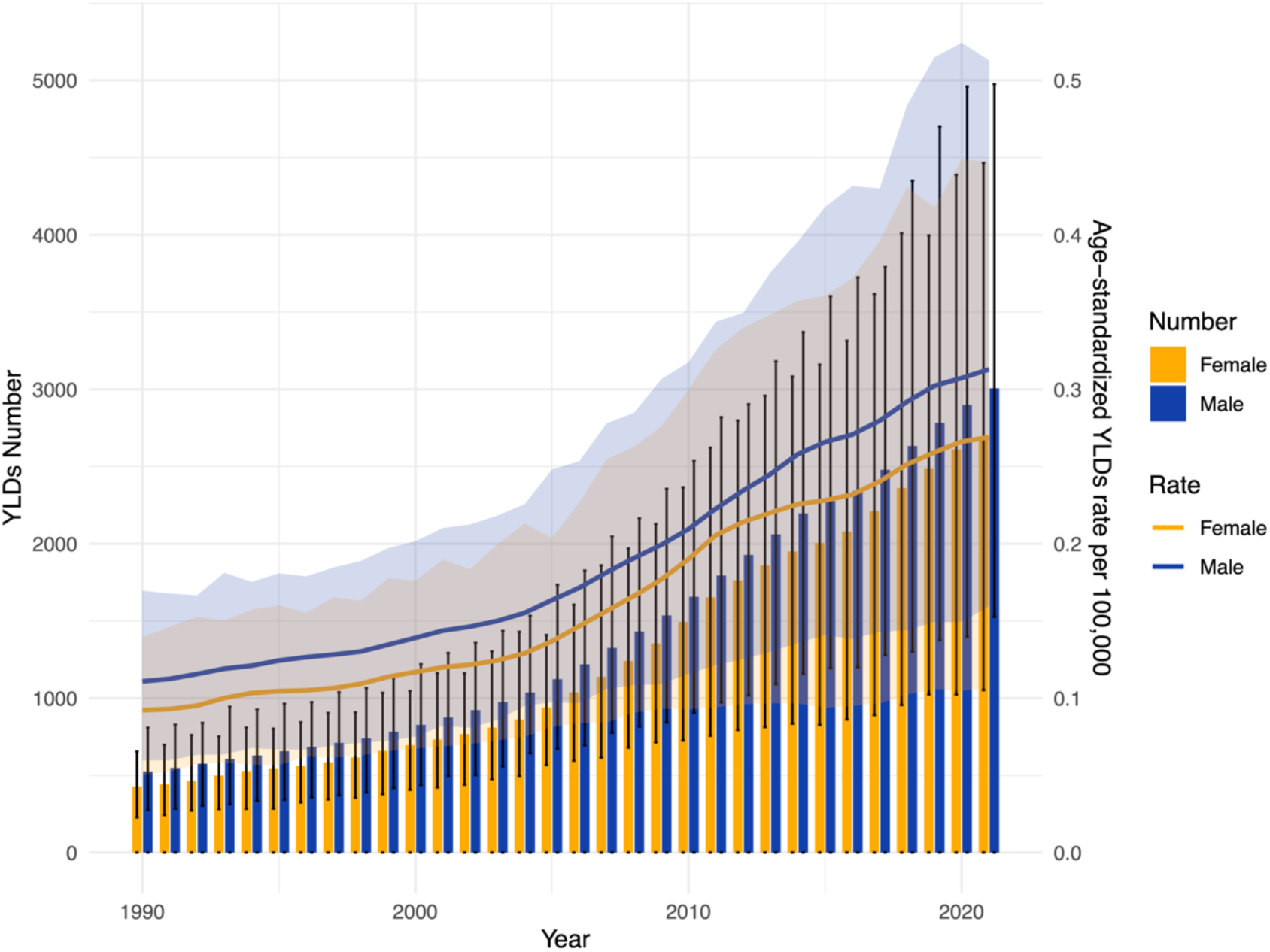
Trends in the YLDs number and age-standardized YLDs rate of MSM by gender in China, from 1990 to 2021.

**Supplementary figure 3.**
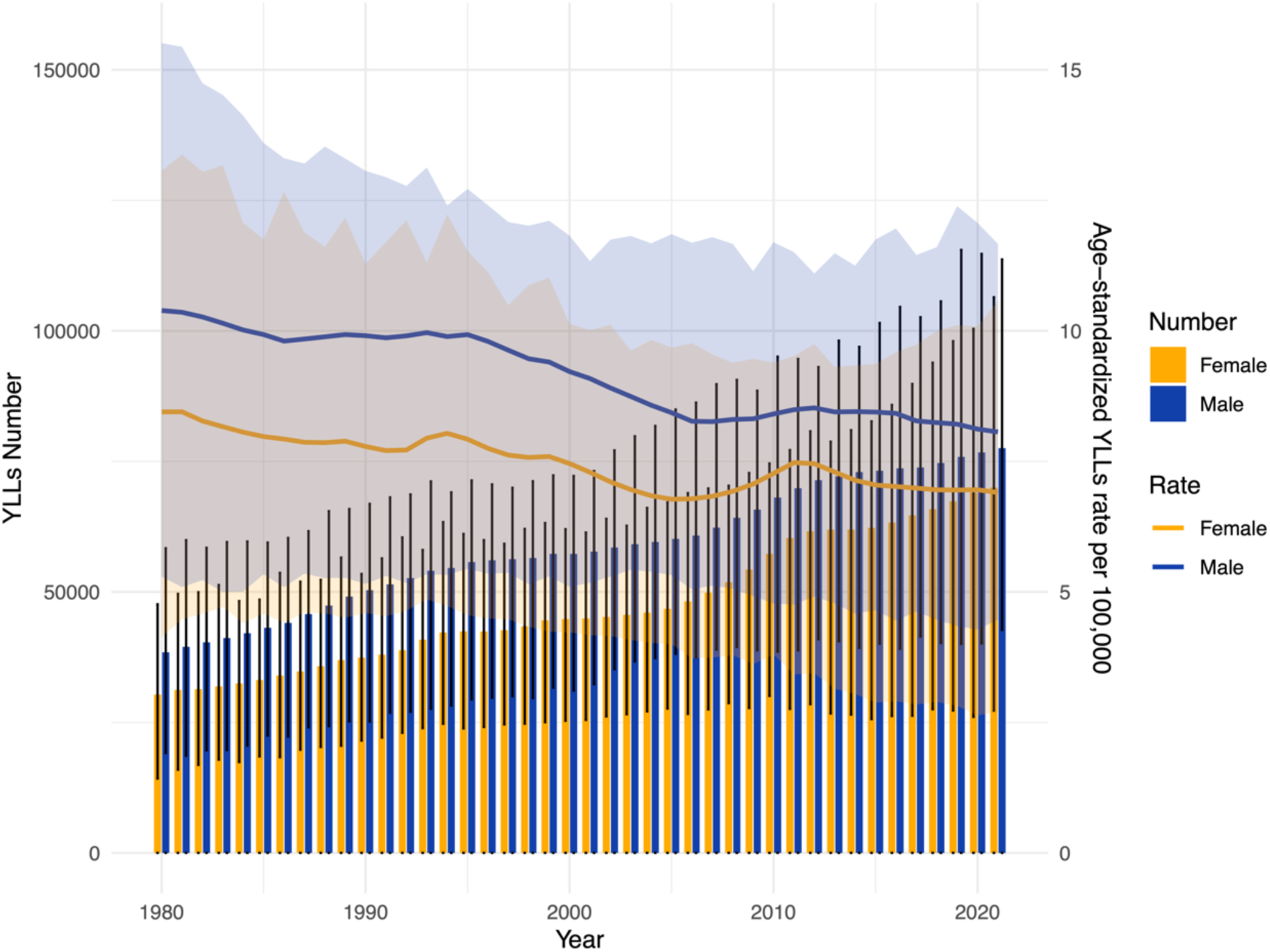
Trends in the YLLs number and age-standardized YLLs rate of MSM by gender in China, from 1990 to 2021.

**Supplementary figure 4.**
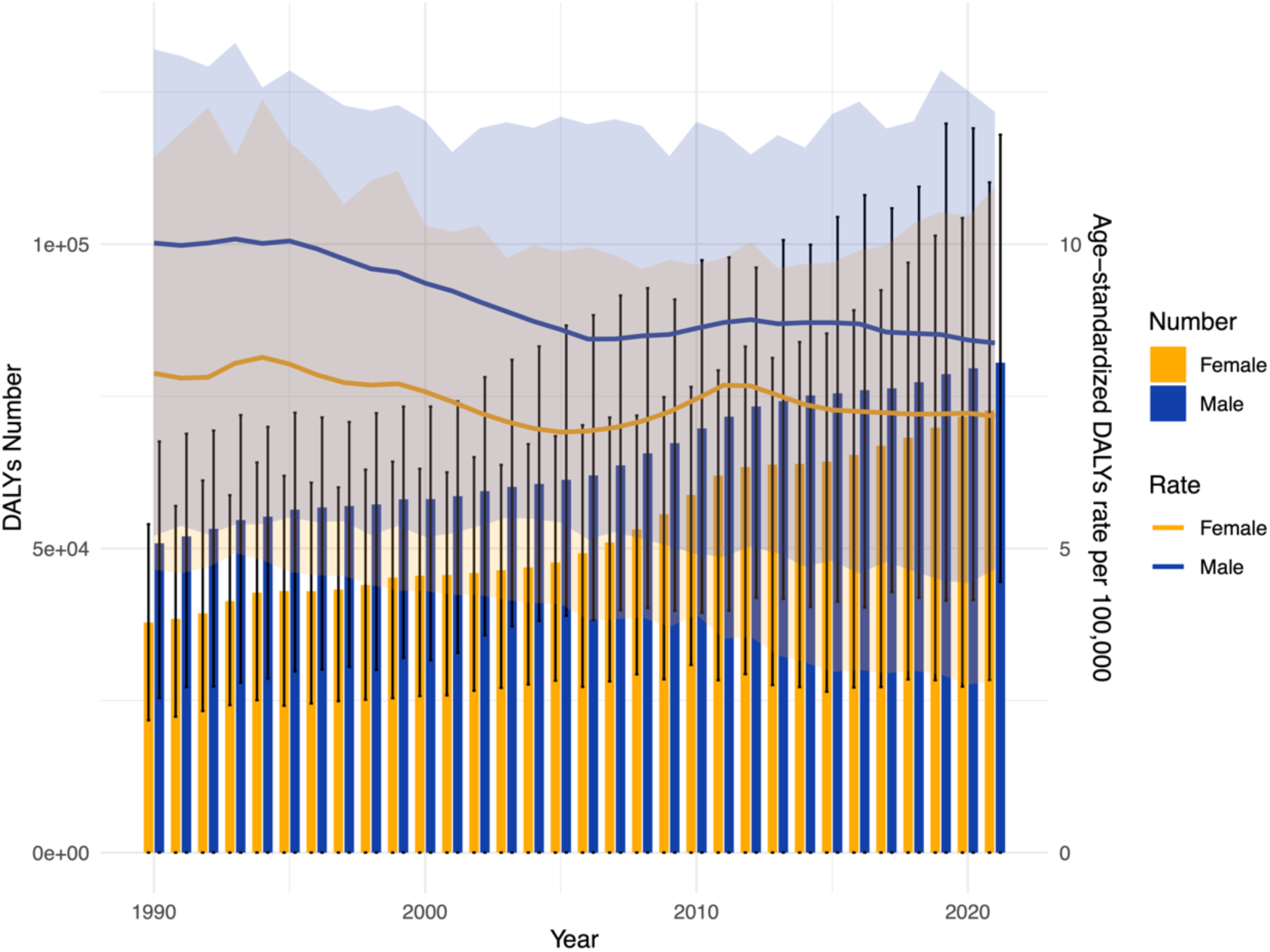
Trends in the DALYs number and age-standardized DALYs rate of MSM by gender in China, from 1990 to 2021.

**Supplementary figure 5.**
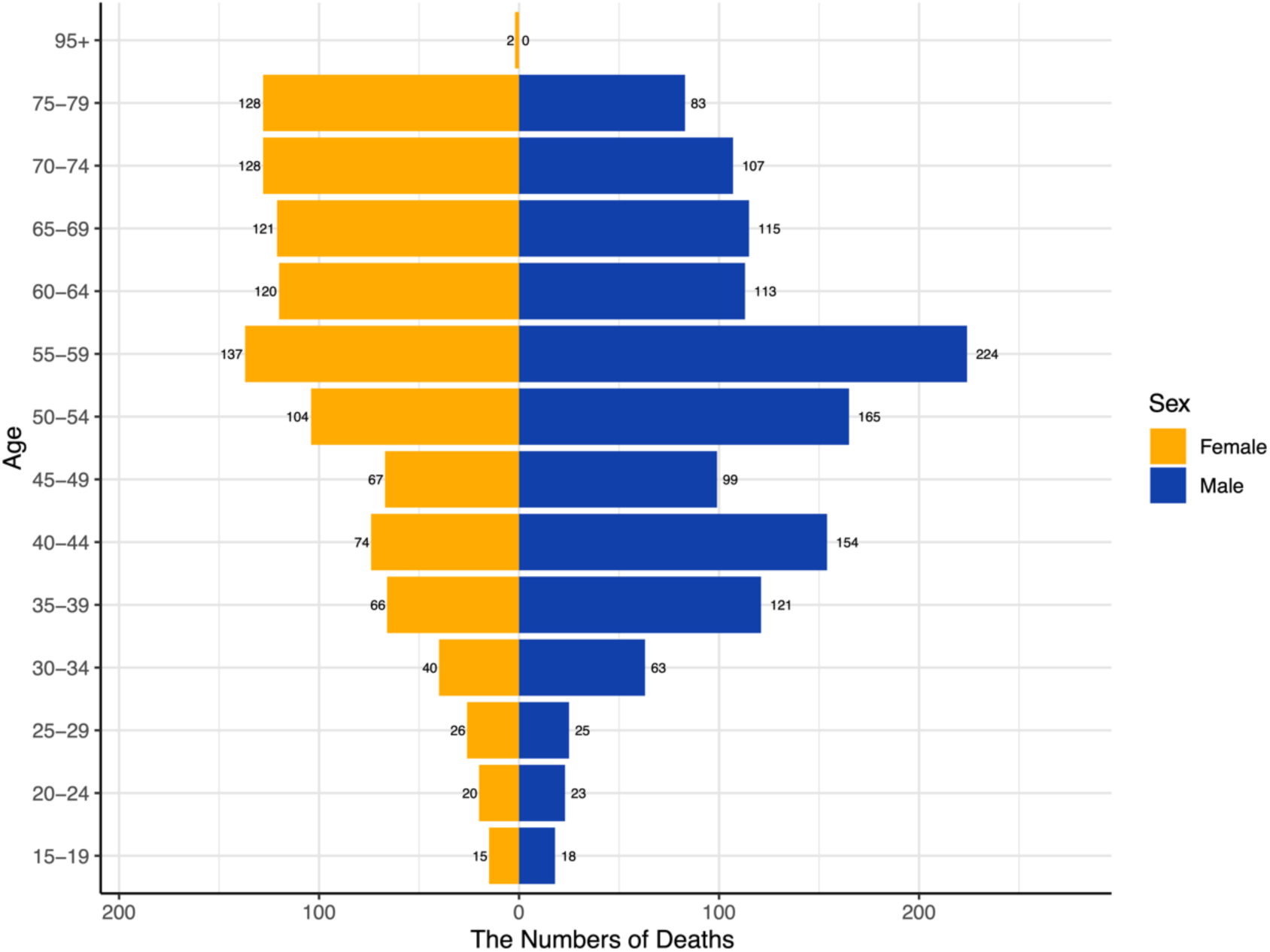
Deaths of new cases among MSM by age group and sex in 1990.

**Supplementary figure 6.**
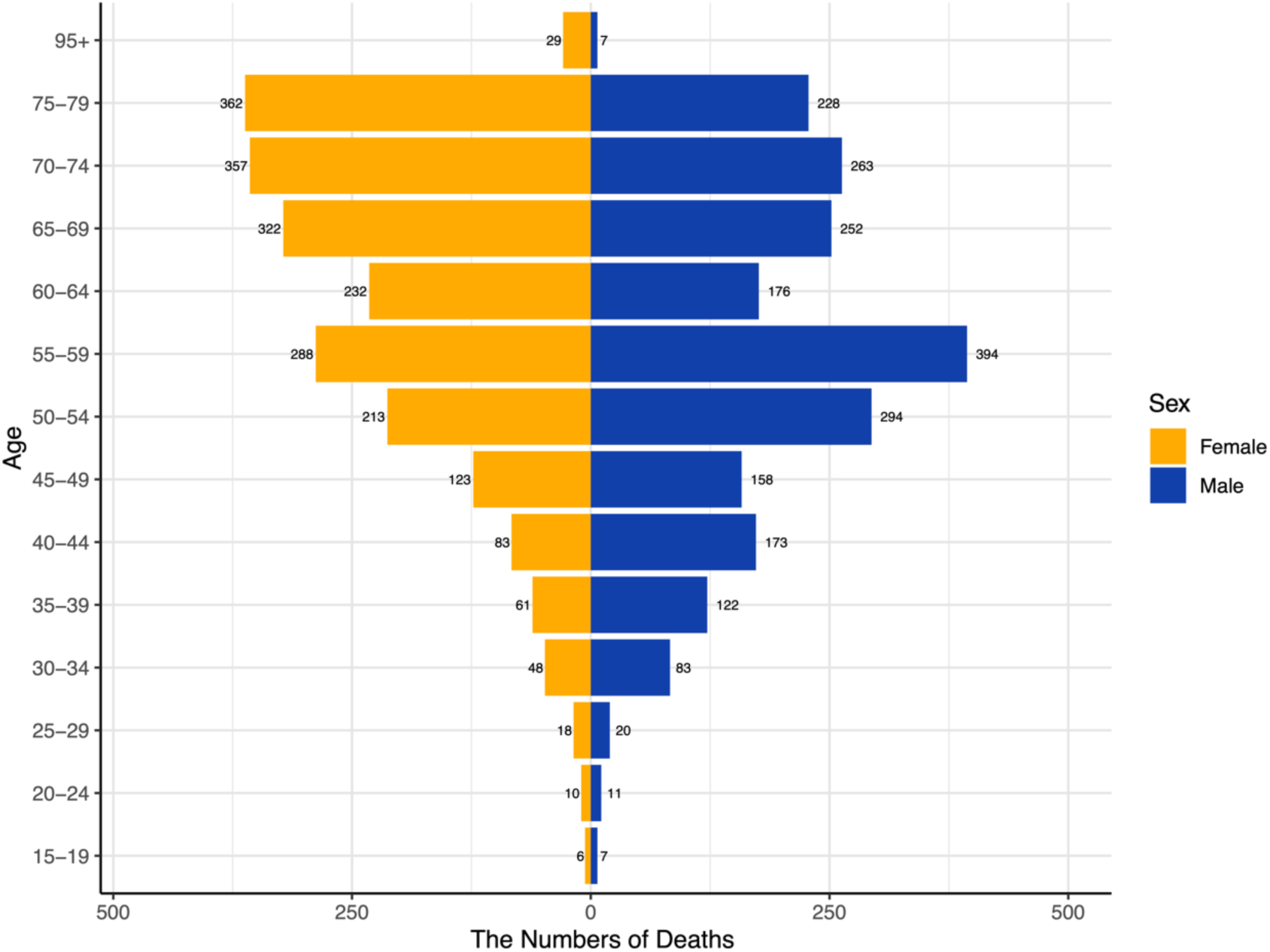
Deaths of new cases among MSM by age group and sex in 2021.

## Notes

### Competing Interest Statement

The authors have declared no competing interest.

## Citation

[1] A. B, Q. Mi, G. S, Malignant Melanoma: Skin Cancer-Diagnosis, Prevention, and Treatment, Critical Reviews in Eukaryotic Gene Expression 30 (2020). 10.1615/CritRevEukaryotGeneExpr.2020028454.

[2] C. Pp, P. V, M. R, The journey from melanocytes to melanoma, Nature Reviews. Cancer 23 (2023). 10.1038/s41568-023-00565-7.

[3] J. Tímár, A. Ladányi, Molecular Pathology of Skin Melanoma: Epidemiology, Differential Diagnostics, Prognosis and Therapy Prediction, Int J Mol Sci 23 (2022) 5384. 10.3390/ijms23105384.

[4] M.A. Kreher, S. Konda, M.M.B. Noland, M.I. Longo, R. Valdes-Rodriguez, Risk of melanoma and nonmelanoma skin cancer with immunosuppressants, part II: Methotrexate, alkylating agents, biologics, and small molecule inhibitors, J Am Acad Dermatol 88 (2023) 534–542. 10.1016/j.jaad.2022.11.043.

[5] N. Turner, O. Ware, M. Bosenberg, Genetics of metastasis: melanoma and other cancers, Clin Exp Metastasis 35 (2018) 379–391. 10.1007/s10585-018-9893-y.

[6] T. H, O. Jm, C. Km, B. Jd, T. Sc, B. Js, C. Mm, S. Aj, T. C, B. R, B. Ws, Early detection of melanoma: reviewing the ABCDEs, Journal of the American Academy of Dermatology 72 (2015). 10.1016/j.jaad.2015.01.025.

[7] M.E. Klapperich, G.M. Bowen, D. Grossman, Current controversies in early-stage melanoma: Questions on management and surveillance, J Am Acad Dermatol 80 (2019) 15–25. 10.1016/j.jaad.2018.03.054.

[8] S.N. Pavri, J. Clune, S. Ariyan, D. Narayan, Malignant Melanoma: Beyond the Basics, Plast Reconstr Surg 138 (2016) 330e–340e. 10.1097/PRS.0000000000002367.

[9] Z.N. Willsmore, B.G.T. Coumbe, S. Crescioli, S. Reci, A. Gupta, R.J. Harris, A. Chenoweth, J. Chauhan, H.J. Bax, A. McCraw, A. Cheung, G. Osborn, R.M. Hoffmann, M. Nakamura, R. Laddach, J.L.C. Geh, A. MacKenzie-Ross, C. Healy, S. Tsoka, J.F. Spicer, D.H. Josephs, S. Papa, K.E. Lacy, S.N. Karagiannis, Combined anti-PD-1 and anti-CTLA-4 checkpoint blockade: Treatment of melanoma and immune mechanisms of action, Eur J Immunol 51 (2021) 544–556. 10.1002/eji.202048747.

[10] L. Tagliaferri, V. Lancellotta, B. Fionda, M. Mangoni, C. Casà, A. Di Stefani, M.M. Pagliara, A. D’Aviero, G. Schinzari, S. Chiesa, C. Mazzarella, S. Manfrida, G.F. Colloca, F. Marazzi, A.G. Morganti, M.A. Blasi, K. Peris, G. Tortora, V. Valentini, Immunotherapy and radiotherapy in melanoma: a multidisciplinary comprehensive review, Hum Vaccin Immunother 18 (2022) 1903827. 10.1080/21645515.2021.1903827.

[11] L.B. Jilaveanu, S.A. Aziz, H.M. Kluger, Chemotherapy and biologic therapies for melanoma: do they work?, Clin Dermatol 27 (2009) 614–625. 10.1016/j.clindermatol.2008.09.020.

[12] C. Li, L. Kuai, R. Cui, X. Miao, Melanogenesis and the Targeted Therapy of Melanoma, Biomolecules 12 (2022) 1874. 10.3390/biom12121874.

[13] R. Pampena, V. Piccolo, M. Muscianese, A. Kyrgidis, M. Lai, T. Russo, G. Briatico, E.V. Di Brizzi, G. Cascone, S. Pellerone, C. Longo, E. Moscarella, G. Argenziano, Melanoma in children: A systematic review and individual patient meta-analysis, J Eur Acad Dermatol Venereol 37 (2023) 1758–1776. 10.1111/jdv.19220.

[14] https://www.csco.org.cn/cn/index.aspx

[15] A.J. Ferrari, D.F, et al., Global incidence, prevalence, years lived with disability (YLDs), disability-adjusted life-years (DALYs), and healthy life expectancy (HALE) for 371 diseases and injuries in 204 countries and territories and 811 subnational locations, 1990–2021: a systematic analysis for the Global Burden of Disease Study 2021, The Lancet 403 (2024) 2133–2161. 10.1016/S0140-6736(24)00757-8.

[16] Y. Kong, P. Xing, P. Huai, F. Zhang, The burden of skin diseases in China: global burden of Disease Study 2019, Arch Dermatol Res 316 (2024) 277. 10.1007/s00403-024-03046-5.

[17] T. Xu, W. Dong, J. Liu, P. Yin, Z. Wang, L. Zhang, M. Zhou, Disease burden of Parkinson’s disease in China and its provinces from 1990 to 2021: findings from the global burden of disease study 2021, The Lancet Regional Health – Western Pacific 46 (2024). 10.1016/j.lanwpc.2024.101078.

[18] C. Kim, S.C. You, J.M. Reps, J.Y. Cheong, R.W. Park, Machine-learning model to predict the cause of death using a stacking ensemble method for observational data, J Am Med Inform Assoc 28 (2021) 1098–1107. 10.1093/jamia/ocaa277.

[19] Q. H, C. S, X. R, Cancer incidence, mortality, and burden in China: a time-trend analysis and comparison with the United States and United Kingdom based on the global epidemiological data released in 2020, Cancer Communications (London, England) 41 (2021). 10.1002/cac2.12197.

[20] D.G. P, Standardization and decomposition of rates from cross-classified data, Genus 50 (1994). https://pubmed.ncbi.nlm.nih.gov/12319256/ (accessed October 12, 2024).

[21] X. Cheng, Y. Yang, D.C. Schwebel, Z. Liu, L. Li, P. Cheng, P. Ning, G. Hu, Population ageing and mortality during 1990-2017: A global decomposition analysis, PLoS Med 17 (2020) e1003138. 10.1371/journal.pmed.1003138.

[22] B. Wagner, K. Cleland, Using autoregressive integrated moving average models for time series analysis of observational data, BMJ 383 (2023) 2739. 10.1136/bmj.p2739.

[23] L. Y, N. Y, S. B, S. Y, S. N, F. Y, D. X, Temporal trends in prevalence and mortality for chronic kidney disease in China from 1990 to 2019: an analysis of the Global Burden of Disease Study 2019, Clinical Kidney Journal 16 (2022). 10.1093/ckj/sfac218.

[24] T. Wu, E. Hu, S. Xu, M. Chen, P. Guo, Z. Dai, T. Feng, L. Zhou, W. Tang, L. Zhan, X. Fu, S. Liu, X. Bo, G. Yu, clusterProfiler 4.0: A universal enrichment tool for interpreting omics data, Innovation (Camb) 2 (2021) 100141. 10.1016/j.xinn.2021.100141.

[25] Y. Zhang, S.M. Ostrowski, D.E. Fisher, Nevi and Melanoma, Hematol Oncol Clin North Am 38 (2024) 939–952. 10.1016/j.hoc.2024.05.005.

[26] P. Chauhan, R. Jindal, E. Errichetti, Dermoscopy of skin parasitoses, bites and stings: a systematic review of the literature, J Eur Acad Dermatol Venereol 36 (2022) 1722–1734. 10.1111/jdv.18352.

[27] N.G. Marghoob, K. Liopyris, N. Jaimes, Dermoscopy: A Review of the Structures That Facilitate Melanoma Detection, J Am Osteopath Assoc 119 (2019) 380–390. 10.7556/jaoa.2019.067.

[28] D. J, D. Jj, C. N, F. di R. L, M. Rn, T. Dr, W. Ky, A. Rb, A. R, F. M, B. Se, G. Mj, T. Y, D. C, G. K, W. Fm, W. Hc, Dermoscopy, with and without visual inspection, for diagnosing melanoma in adults, The Cochrane Database of Systematic Reviews 12 (2018). 10.1002/14651858.CD011902.pub2.

[29] K H., Z J., Y L., Z W., X W., W Z., M C., Y Z., K Z., Y L., N Y., S L., X H., J S., M Y., B Q., X W., X C., S Z., The Classification of Six Common Skin Diseases Based on Xiangya-Derm: Development of a Chinese Database for Artificial Intelligence, Journal of medical Internet research 23 (2021). 10.2196/26025.

[30] Shen X., Yu R.-X., Shen C.-B., Li C.-X., Jing Y., Zheng Y.-J., Wang Z.-Y., Xue K., Xu F., Yu J.-B., Meng R.-S., Cui Y., Dermoscopy in China: current status and future prospective, Chinese Medical Journal 132 (2019) 2096. 10.1097/CM9.0000000000000396.

[31] T. Wu, X. Wang, S. Zhao, Y. Xiao, M. Shen, X. Han, X. Chen, J. Su, Socioeconomic Determinants of Melanoma-Related Health Literacy and Attitudes Among College Students in China: A Population-Based Cross-Sectional Study, Front. Public Health 9 (2021). 10.3389/fpubh.2021.743368.

[32] L S., Y K., X X., Kt F., X S., C C., Z C., S L., L M., J G., Prevalence of BRAF V600E mutation in Chinese melanoma patients: large scale analysis of BRAF and NRAS mutations in a 432-case cohort, European journal of cancer (Oxford, England : 1990) 48 (2012). 10.1016/j.ejca.2011.06.056.

[33] X. Bai, K.T. Flaherty, Targeted and immunotherapies in BRAF mutant melanoma: where we stand and what to expect, Br J Dermatol 185 (2021) 253–262. 10.1111/bjd.19394.

[34] E. Chrysanthou, E. Sehovic, P. Ostano, G. Chiorino, Comprehensive Gene Expression Analysis to Identify Differences and Similarities between Sex- and Stage-Stratified Melanoma Samples, Cells 11 (2022) 1099. 10.3390/cells11071099.

[35] C.M. Olsen, J.F. Thompson, N. Pandeya, D.C. Whiteman, Evaluation of Sex-Specific Incidence of Melanoma, JAMA Dermatology 156 (2020) 553–560. 10.1001/jamadermatol.2020.0470.

[36] X. Ke, W. Lin, D. Li, S. Zhao, M. Chen, Y. Xiao, X. Chen, M. Shen, J. Su, Spending and Hospital Stay for Melanoma in Hunan, China, Front. Public Health 10 (2022). 10.3389/fpubh.2022.917119.

[37] Y.-J. Cai, L.-F. Ke, W.-W. Zhang, J.-P. Lu, Y.-P. Chen, Recurrent KRAS, KIT and SF3B1 mutations in melanoma of the female genital tract, BMC Cancer 21 (2021) 677. 10.1186/s12885-021-08427-x.

[38] H. Sm, E. Mr, Phenotype Switching and the Melanoma Microenvironment; Impact on Immunotherapy and Drug Resistance, International Journal of Molecular Sciences 24 (2023). 10.3390/ijms24021601.

[39] R. M, B. A, V. Ic, A. D, F. M, M. P, L. A, de V. M, G. A, Immunotherapy in the Treatment of Metastatic Melanoma: Current Knowledge and Future Directions, Journal of Immunology Research 2020 (2020). 10.1155/2020/9235638.

[40] O. Michielin, A.C.J. van Akkooi, P.A. Ascierto, R. Dummer, U. Keilholz, ESMO Guidelines Committee. Electronic address, Cutaneous melanoma: ESMO Clinical Practice Guidelines for diagnosis, treatment and follow-up†, Ann Oncol 30 (2019) 1884–1901. 10.1093/annonc/mdz411.

[41] Z. Lin, H. Shen, X. Liu, W. Ma, M. Wang, J. Ruan, H. Yu, S. Ma, X. Sun, Naya, Recent advances of artificial intelligence in melanoma clinical practice, Melanoma Res 33 (2023) 454–461. 10.1097/CMR.0000000000000922.

[42] H. F, C. F, D. Ms, G. K, T. A, D. J, E.-T. F, P. Jy, L. M, G. C, G. N, S. J, L. Jh, G. P, J. Js, M. Jj, S. Jv, M. Wh, D.R. Sv, Peroxisome disruption alters lipid metabolism and potentiates antitumor response with MAPK-targeted therapy in melanoma, The Journal of Clinical Investigation 133 (2023). 10.1172/JCI166644.

