## Supplementary figure 1 for "The Burden of Malignant Skin Melanoma in China: From 1990 to 2021 with Estimation to 2036"

| Measure | Percentage change in ASRs per 100,000 people from 1990 to 2021 | | |
|---|---|---|---|
| | Both | Male | Female |
| Incidence | 0.89 (0.15,1.58) | 0.84 (0.19,1.86) | 0.96 (-0.1,1.95) |
| Prevalence | 5.18 (2.7,7.77) | 5.17 (2.92,9.25) | 5.2 (1.83,8.67) |
| Deaths | -0.11 (-0.46,0.19) | -0.17 (-0.44,0.2) | -0.05 (-0.56,0.41) |
| YLDs | 1.86 (0.72,3.12) | 1.82 (0.72,3.48) | 1.92 (0.31,3.62) |
| YLLs | -0.16 (-0.49,0.15) | -0.19 (-0.48,0.25) | -0.11 (-0.61,0.36) |
| DALYs | -0.13 (-0.48,0.19) | -0.16 (-0.46,0.3) | -0.09 (-0.59,0.4) |

Supplementary table 1. Incidence, prevalence, deaths, YLDs, YLLs and DALYs percentage change for malignant skin melanoma from 1990 to 2021. ASRs, age-standardized rates; YLLs, years of life lost; YLDs, years of healthy life lost due to disability; DALYs, disability adjusted life year. 95 % UI=95 % uncertainty intervals.

| ASIR_male | Year | Value | Type | ASIR_female | Year | Value | Type |
|---|---|---|---|---|---|---|---|
| 1 | 1990 | 0.39637977 | Actual | 1 | 1990 | 0.32607526 | Actual |
| 2 | 1991 | 0.39906111 | Actual | 2 | 1991 | 0.32631697 | Actual |
| 3 | 1992 | 0.40761603 | Actual | 3 | 1992 | 0.33168652 | Actual |
| 4 | 1993 | 0.41680393 | Actual | 4 | 1993 | 0.3465588 | Actual |
| 5 | 1994 | 0.42090032 | Actual | 5 | 1994 | 0.35541087 | Actual |
| 6 | 1995 | 0.43035819 | Actual | 6 | 1995 | 0.35663991 | Actual |
| 7 | 1996 | 0.43242353 | Actual | 7 | 1996 | 0.35459196 | Actual |
| 8 | 1997 | 0.43190274 | Actual | 8 | 1997 | 0.35430638 | Actual |
| 9 | 1998 | 0.43219382 | Actual | 9 | 1998 | 0.35797341 | Actual |
| 10 | 1999 | 0.43806019 | Actual | 10 | 1999 | 0.36491742 | Actual |
| 11 | 2000 | 0.43921216 | Actual | 11 | 2000 | 0.36505065 | Actual |
| 12 | 2001 | 0.4425802 | Actual | 12 | 2001 | 0.36464092 | Actual |
| 13 | 2002 | 0.44144946 | Actual | 13 | 2002 | 0.36196178 | Actual |
| 14 | 2003 | 0.44525966 | Actual | 14 | 2003 | 0.36324967 | Actual |
| 15 | 2004 | 0.4552444 | Actual | 15 | 2004 | 0.37132105 | Actual |
| 16 | 2005 | 0.47150318 | Actual | 16 | 2005 | 0.38768777 | Actual |
| 17 | 2006 | 0.48490829 | Actual | 17 | 2006 | 0.40778915 | Actual |
| 18 | 2007 | 0.50538398 | Actual | 18 | 2007 | 0.42866373 | Actual |
| 19 | 2008 | 0.52273149 | Actual | 19 | 2008 | 0.4489779 | Actual |
| 20 | 2009 | 0.53399744 | Actual | 20 | 2009 | 0.46973255 | Actual |
| 21 | 2010 | 0.55218457 | Actual | 21 | 2010 | 0.49729255 | Actual |
| 22 | 2011 | 0.57980242 | Actual | 22 | 2011 | 0.53131807 | Actual |
| 23 | 2012 | 0.60454434 | Actual | 23 | 2012 | 0.54775046 | Actual |
| 24 | 2013 | 0.62017844 | Actual | 24 | 2013 | 0.55387432 | Actual |
| 25 | 2014 | 0.64089708 | Actual | 25 | 2014 | 0.55905014 | Actual |
| 26 | 2015 | 0.65444269 | Actual | 26 | 2015 | 0.56110303 | Actual |
| 27 | 2016 | 0.66326212 | Actual | 27 | 2016 | 0.56760864 | Actual |
| 28 | 2017 | 0.67272349 | Actual | 28 | 2017 | 0.58275128 | Actual |
| 29 | 2018 | 0.69378965 | Actual | 29 | 2018 | 0.60124314 | Actual |
| 30 | 2019 | 0.71228327 | Actual | 30 | 2019 | 0.61881408 | Actual |
| 31 | 2020 | 0.72040071 | Actual | 31 | 2020 | 0.63110449 | Actual |
| 32 | 2021 | 0.73047775 | Actual | 32 | 2021 | 0.63763828 | Actual |
| 33 | 2022 | 0.7405548 | Forecast | 33 | 2022 | 0.64245623 | Forecast |
| 34 | 2023 | 0.75063185 | Forecast | 34 | 2023 | 0.64727419 | Forecast |
| 35 | 2024 | 0.76070889 | Forecast | 35 | 2024 | 0.65209214 | Forecast |
| 36 | 2025 | 0.77078594 | Forecast | 36 | 2025 | 0.6569101 | Forecast |
| 37 | 2026 | 0.78086298 | Forecast | 37 | 2026 | 0.66172806 | Forecast |
| 38 | 2027 | 0.79094003 | Forecast | 38 | 2027 | 0.66654601 | Forecast |
| 39 | 2028 | 0.80101708 | Forecast | 39 | 2028 | 0.67136397 | Forecast |
| 40 | 2029 | 0.81109412 | Forecast | 40 | 2029 | 0.67618192 | Forecast |
| 41 | 2030 | 0.82117117 | Forecast | 41 | 2030 | 0.68099988 | Forecast |

| | | | | | | | |
|---|---|---|---|---|---|---|---|
| 42 | 2031 | 0.83124821 | Forecast | 42 | 2031 | 0.68581783 | Forecast |
| 43 | 2032 | 0.84132526 | Forecast | 43 | 2032 | 0.69063579 | Forecast |
| 44 | 2033 | 0.85140231 | Forecast | 44 | 2033 | 0.69545375 | Forecast |
| 45 | 2034 | 0.86147935 | Forecast | 45 | 2034 | 0.7002717 | Forecast |
| 46 | 2035 | 0.8715564 | Forecast | 46 | 2035 | 0.70508966 | Forecast |
| 47 | 2036 | 0.88163344 | Forecast | 47 | 2036 | 0.70990761 | Forecast |

Supplementary table 2. ARIMA forecast data of ASIR by gender.

| ASDR_male | Year | Value | Type | ASDR_female | Year | Value | Type |
|---|---|---|---|---|---|---|---|
| 1 | 1990 | 0.33375351 | Actual | 1 | 1990 | 0.28141521 | Actual |
| 2 | 1991 | 0.33275469 | Actual | 2 | 1991 | 0.27910927 | Actual |
| 3 | 1992 | 0.33720745 | Actual | 3 | 1992 | 0.28156883 | Actual |
| 4 | 1993 | 0.3415465 | Actual | 4 | 1993 | 0.29134016 | Actual |
| 5 | 1994 | 0.34106929 | Actual | 5 | 1994 | 0.29555417 | Actual |
| 6 | 1995 | 0.34540331 | Actual | 6 | 1995 | 0.29350376 | Actual |
| 7 | 1996 | 0.34227768 | Actual | 7 | 1996 | 0.28800792 | Actual |
| 8 | 1997 | 0.33695822 | Actual | 8 | 1997 | 0.28361252 | Actual |
| 9 | 1998 | 0.3321585 | Actual | 9 | 1998 | 0.28224899 | Actual |
| 10 | 1999 | 0.33244753 | Actual | 10 | 1999 | 0.28383961 | Actual |
| 11 | 2000 | 0.32841989 | Actual | 11 | 2000 | 0.2795969 | Actual |
| 12 | 2001 | 0.32644046 | Actual | 12 | 2001 | 0.27533785 | Actual |
| 13 | 2002 | 0.31803169 | Actual | 13 | 2002 | 0.26694725 | Actual |
| 14 | 2003 | 0.31271294 | Actual | 14 | 2003 | 0.26006444 | Actual |
| 15 | 2004 | 0.308696 | Actual | 15 | 2004 | 0.25731426 | Actual |
| 16 | 2005 | 0.30428482 | Actual | 16 | 2005 | 0.25643622 | Actual |
| 17 | 2006 | 0.2967471 | Actual | 17 | 2006 | 0.25664822 | Actual |
| 18 | 2007 | 0.29653975 | Actual | 18 | 2007 | 0.25984814 | Actual |
| 19 | 2008 | 0.29572686 | Actual | 19 | 2008 | 0.26342897 | Actual |
| 20 | 2009 | 0.29125968 | Actual | 20 | 2009 | 0.26719641 | Actual |
| 21 | 2010 | 0.29102032 | Actual | 21 | 2010 | 0.27482495 | Actual |
| 22 | 2011 | 0.29580909 | Actual | 22 | 2011 | 0.28499076 | Actual |
| 23 | 2012 | 0.2978419 | Actual | 23 | 2012 | 0.28418399 | Actual |
| 24 | 2013 | 0.2913065 | Actual | 24 | 2013 | 0.27479843 | Actual |
| 25 | 2014 | 0.28843488 | Actual | 25 | 2014 | 0.26707943 | Actual |
| 26 | 2015 | 0.2922376 | Actual | 26 | 2015 | 0.26685125 | Actual |
| 27 | 2016 | 0.29323908 | Actual | 27 | 2016 | 0.26851454 | Actual |
| 28 | 2017 | 0.28326293 | Actual | 28 | 2017 | 0.26614199 | Actual |
| 29 | 2018 | 0.28104488 | Actual | 29 | 2018 | 0.26575859 | Actual |
| 30 | 2019 | 0.28201239 | Actual | 30 | 2019 | 0.2682412 | Actual |
| 31 | 2020 | 0.27886151 | Actual | 31 | 2020 | 0.26923605 | Actual |
| 32 | 2021 | 0.27724048 | Actual | 32 | 2021 | 0.26697666 | Actual |
| 33 | 2022 | 0.27573845 | Forecast | 33 | 2022 | 0.26439747 | Forecast |
| 34 | 2023 | 0.27391954 | Forecast | 34 | 2023 | 0.26325892 | Forecast |
| 35 | 2024 | 0.27210064 | Forecast | 35 | 2024 | 0.2635976 | Forecast |
| 36 | 2025 | 0.27028173 | Forecast | 36 | 2025 | 0.26451046 | Forecast |
| 37 | 2026 | 0.26846283 | Forecast | 37 | 2026 | 0.26515861 | Forecast |
| 38 | 2027 | 0.26664393 | Forecast | 38 | 2027 | 0.2652612 | Forecast |
| 39 | 2028 | 0.26482502 | Forecast | 39 | 2028 | 0.26500957 | Forecast |
| 40 | 2029 | 0.26300612 | Forecast | 40 | 2029 | 0.26472668 | Forecast |
| 41 | 2030 | 0.26118721 | Forecast | 41 | 2030 | 0.26460384 | Forecast |

| | | | | | | | |
|---|---|---|---|---|---|---|---|
| 42 | 2031 | 0.25936831 | Forecast | 42 | 2031 | 0.26464283 | Forecast |
| 43 | 2032 | 0.2575494 | Forecast | 43 | 2032 | 0.26474354 | Forecast |
| 44 | 2033 | 0.2557305 | Forecast | 44 | 2033 | 0.26481419 | Forecast |
| 45 | 2034 | 0.2539116 | Forecast | 45 | 2034 | 0.26482472 | Forecast |
| 46 | 2035 | 0.25209269 | Forecast | 46 | 2035 | 0.26479671 | Forecast |
| 47 | 2036 | 0.25027379 | Forecast | 47 | 2036 | 0.26476569 | Forecast |

Supplementary table 3. ARIMA forecast data of ASDR by gender.

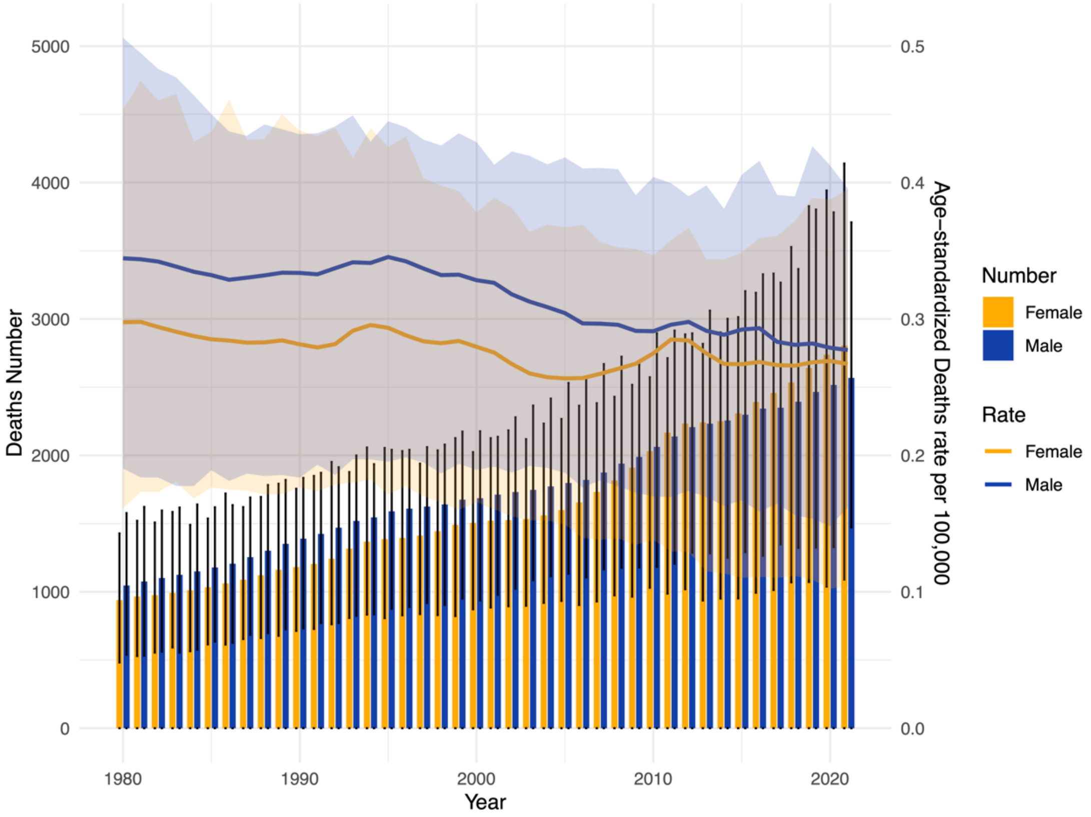


Supplementary figure 1. Trends in the deaths number and age-standardized deaths rate (ASDR) of MSM by gender in China, from 1990 to 2021.

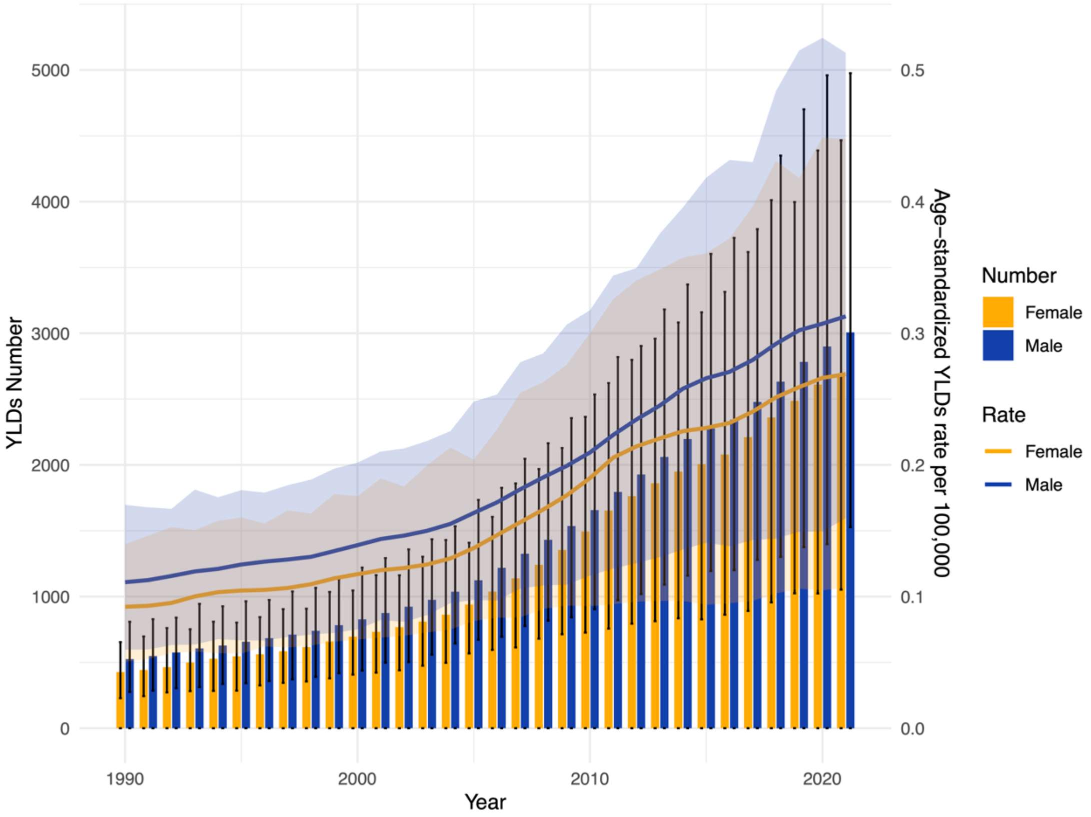


Supplementary figure 2. Trends in the YLDs number and age-standardized YLDs rate of MSM by gender in China, from 1990 to 2021.

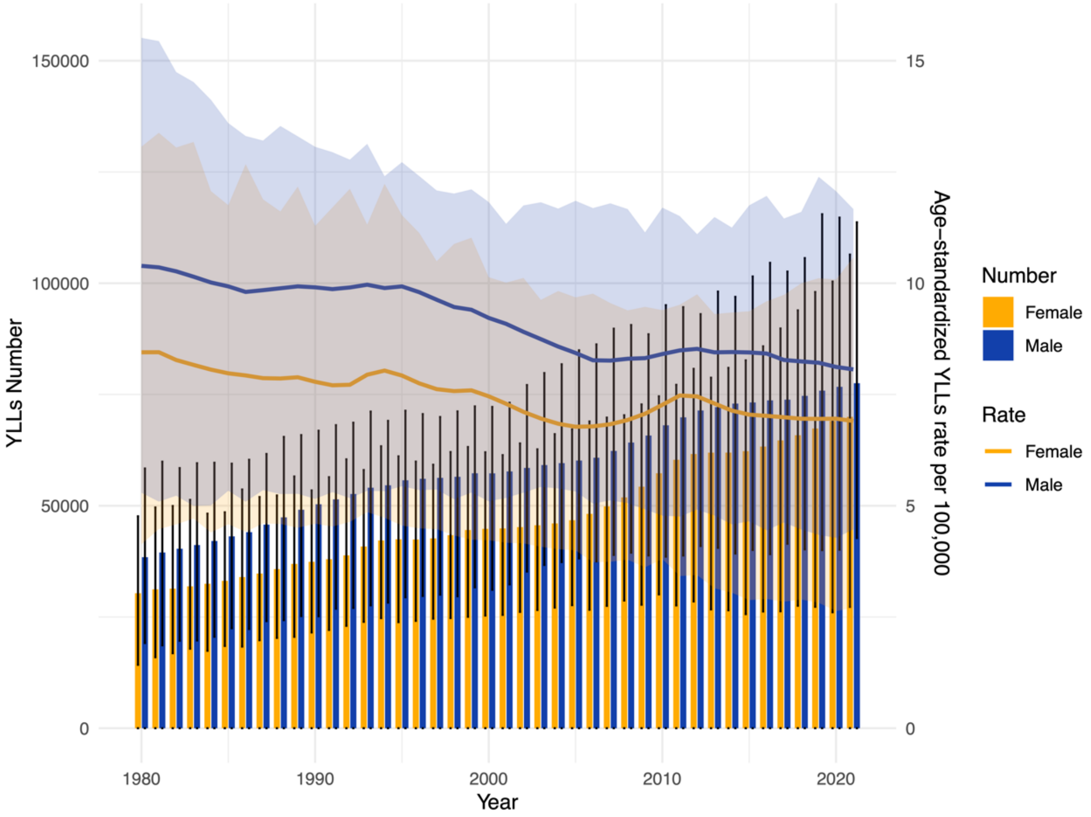


Supplementary figure 3. Trends in the YLLs number and age-standardized YLLs rate of MSM by gender in China, from 1990 to 2021.

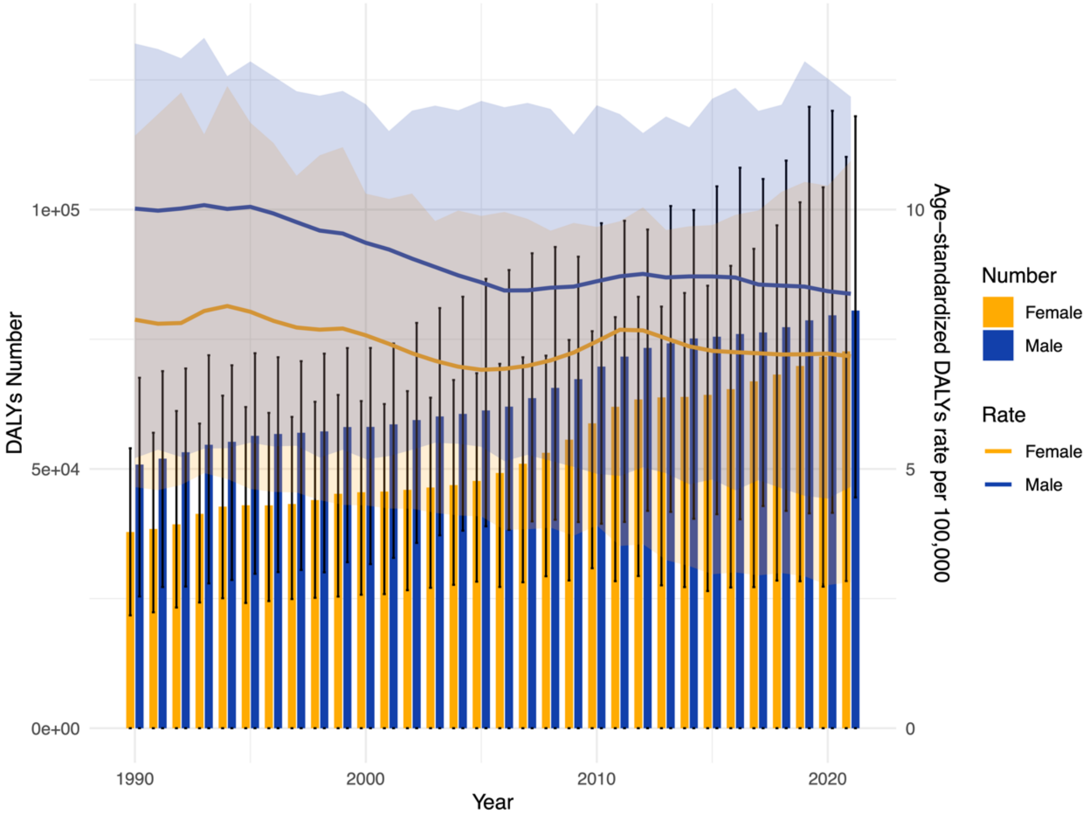


Supplementary figure 4. Trends in the DALYs number and age-standardized DALYs rate of MSM by gender in China, from 1990 to 2021.

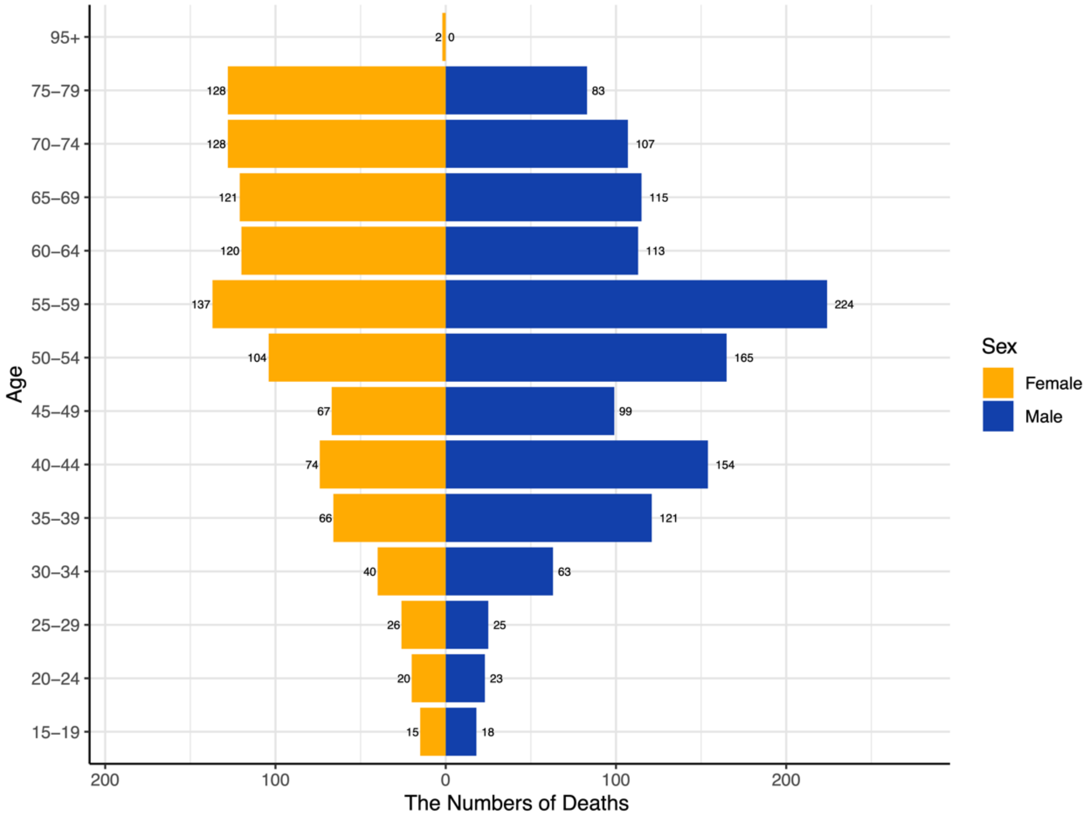


Supplementary figure 5. Deaths of new cases among MSM by age group and sex in 1990.

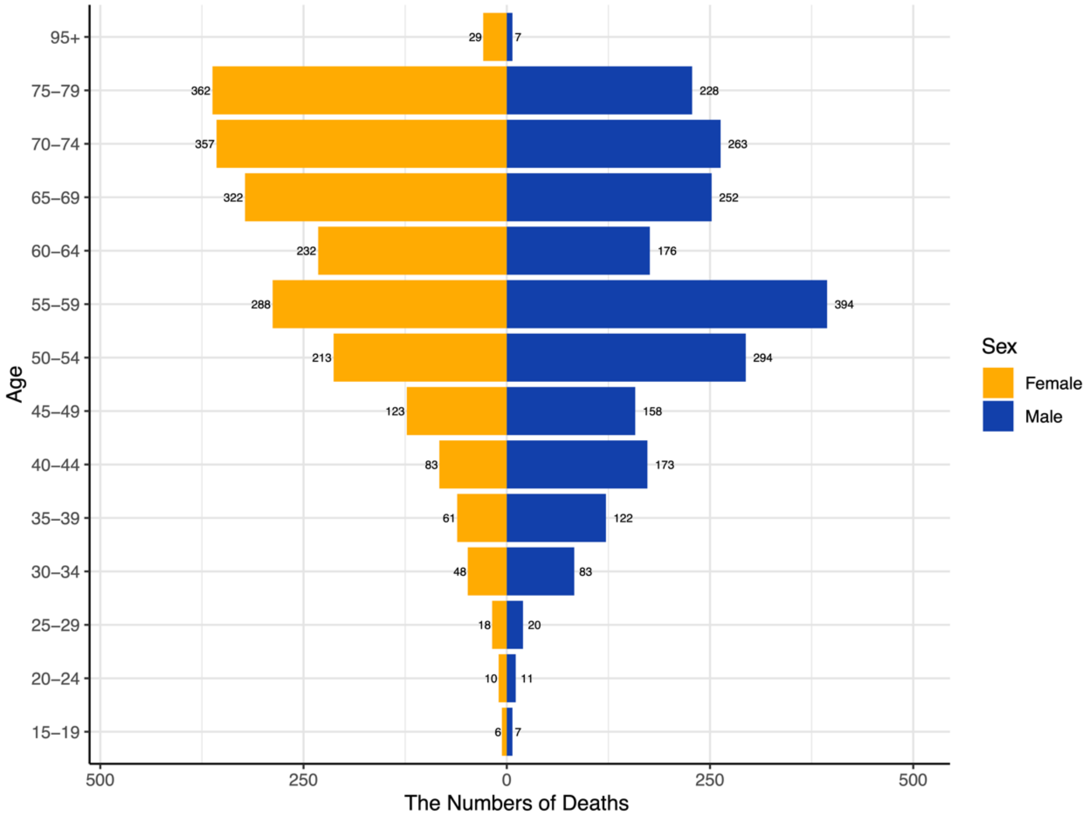


Supplementary figure 6. Deaths of new cases among MSM by age group and sex in 2021.
